# Elevation in Plasma p-tau217 Concentrations Among Blood Donors: An Exploratory Cross-Sectional Study

**DOI:** 10.64898/2026.09.23.26363733

**Authors:** Xiang Li, Weiting Zhang, Ruiwei Wang, Hlaing Tint, Shengyang Fu, Qimin Quan, Feng Liang, Cindy Tint, Xiaoyi Yuan, YaFeng Liang, Yandong Jiang, Wei Cao, Yuanlin Dong, Yuansong Zhao, Sepideh Saroukani, Yiying Zhang, Louise McCullough, Holger K. Eltzschig, Alparslan Turan, Daniel I. Sessler, Thomas K. Karikari, Zhongcong Xie

**Affiliations:** Department of Anesthesiology, Critical Care and Pain Medicine, Institute of Perioperative Medicine, McGovern Medical School, University of Texas Health Science Center at Houston, Houston, TX 77030, USA; Department of Pathology, McGovern Medical School, University of Texas Health Science Center at Houston, Houston, TX 77030, USA; NanoMosaic, Inc., Waltham, MA 02451, USA; Department of Anesthesia, Critical Care and Pain Medicine, Massachusetts General Hospital and Harvard Medical School, Boston, MA 02124, USA; OUTCOMES RESEARCH Consortium®, Houston, TX 77030, USA; Center for Outcomes Research and Department of Anesthesiology, Critical Care and Pain Medicine, McGovern Medical School at UTHealth Houston, Houston, TX 77030, USA; Division of Clinical and Translational Sciences, Department of Internal Medicine, McGovern Medical School, The University of Texas Health Science Center at Houston, Houston, TX 77030, USA; Biostatistics/Epidemiology/Research Design (BERD) component, Center for Clinical and Translational Sciences (CCTS), The University of Texas Health Science Center at Houston, Houston, TX 77030, USA; Department of Neurology, McGovern Medical School, University of Texas Health Science Center at Houston, Houston, TX 77030, USA; Biofluid Biomarker Laboratory, Department of Psychiatry, Western Psychiatric Hospital, Alzheimer’s Disease Research Center, University of Pittsburgh Medical Center, Pittsburgh, PA 15213, USA

## Abstract

**Importance:** Phosphorylated tau at threonine 217 (p-tau217) and brain-derived tau (BD-tau) are increasingly used as blood biomarkers of Alzheimer’s disease (AD) and related neurological disorders. However, the distributions and biological correlates of elevated plasma p-tau217 and BD-tau in plasma from individuals eligible for blood donation remain unclear.

**Objective:** To estimate the proportions of plasma p-tau217 and BD-tau exceeding exploratory study-specific thresholds, and examine associated inflammatory and transcriptomic features.

**Design:** Cross-sectional study.

**Setting:** Hospital blood bank.

**Participants:** 257 de-identified donor samples were obtained between September 2025 and June 2026. Plasma samples from 30 population reference participants and 20 AD patients were used to derive study-specific thresholds. Complementary transcriptomic analyses were performed in 29 fresh whole blood samples.

**Main Outcomes and Measures:** The primary outcome was the proportion of donor samples exceeding the study-specific ROC-derived Youden threshold. The proportion exceeding the prespecified published p-tau217 threshold was the secondary outcome. Other measures included plasma concentrations of BD-tau, tumor necrosis factor α (TNF-α), and interleukin 6 (IL-6); correlations among plasma p-tau217, BD-tau and inflammatory cytokines; and associated transcriptomic alterations.

**Results:** Plasma p-tau217 concentrations exceeded the study-specific ROC-derived Youden threshold in 53 of 256 donor samples (21%; 95% CI, 16%-26%) and the published threshold, used for a secondary comparison, in 99 of 256 samples (39%; 95% CI, 33%-45%). Plasma BD-tau concentrations exceeded the study-specific threshold in 89 of 257 samples (35%; 95% CI, 29%-41%). Plasma p-tau217 concentrations were positively associated with plasma TNF-α (Kendall’s τ = 0.339; *P* < .001) and IL-6 (Kendall’s τ = 0.310; *P* < .001) concentrations. Exploratory transcriptomic analyses identified gene expression patterns associated with p-tau217 concentrations. Measurement using a second p-tau217 antibody showed the similar findings.

**Conclusions and Relevance:** Plasma p-tau217 and BD-tau concentrations exceeded exploratory study-specific thresholds in a subset of blood-donor samples, and higher p-tau217 concentrations were associated with inflammatory biomarkers and exploratory transcriptomic differences. These findings raise the possibility of elevated circulating tau species in some blood donors, but their clinical significance requires confirmation. Whether elevated circulating p-tau217 in donated blood has biological consequences for transfusion recipients is unknown, but deserves investigation.

**Key Points:** **Question:** How commonly are plasma p-tau217 and brain-derived tau concentrations elevated in blood donors, and are these elevations associated with systemic inflammatory and transcriptomic signatures?

**Findings:** In this cross-sectional study of 257 blood-donor samples, 21% of evaluable samples exceeded the study-specific p-tau217 threshold and 35% exceeded the BD-tau threshold. Higher p-tau217 concentrations were associated with inflammatory biomarkers and exploratory differences in gene expression.

**Meaning:** These findings raise the possibility of elevated circulating tau species in blood donor, although confirmation is needed to establish whether these concentrations represent clinically meaningful elevations.

## Introduction

Blood-based biomarkers have improved the diagnosis and monitoring of Alzheimer’s disease (AD) and related neurodegenerative disorders ^1,2^. Among them, plasma phosphorylated tau at threonine 217 (p-tau217) is a useful biomarker of AD and increasingly used both for research and clinical care ^1–5^. Brain-derived tau (BD-tau) is also a potential biomarker of neurodegeneration, neuronal injury, and AD ^6,7^. Consequently, elevated plasma p-tau217 and BD-tau concentrations are often interpreted as indicators of underlying neurodegenerative processes ^5^.

Emerging evidence suggests that circulating tau species may be influenced by factors beyond AD pathology, including kidney dysfunction, non-AD neurologic diseases, and peripheral sources of tau ^6–10^. For example, experimental and clinical studies indicate that tau protein may be released from peripheral tissues ^8,11^ and possibly contributes to inflammatory and immune pathways ^12^. Plasma p-tau217 concentrations vary considerably among individuals ^13^.

While evaluating blood-based biomarkers, we unexpectedly observed elevated plasma p-tau217 concentrations in five donated human plasma samples. Whether such elevations represent isolated analytical findings or are common remains unknown, as is the biological relevance of elevated circulating p-tau217.

We therefore measured plasma p-tau217 and BD-tau concentrations in human plasma samples from blood donors by using ultrasensitive nanoneedle-based assays ^14,15^ with two different antibodies. Given the reported interaction between tau phosphorylation and inflammation ^12^, we examined associations between plasma p-tau217 concentrations and inflammatory cytokines in blood donors. Finally, we determined transcriptomic alterations in fresh human whole blood to explore the potential biological relevance of elevated circulating p-tau. Specifically, we investigated whether plasma p-tau217 concentrations exceed a study-specific threshold in a subset of blood donors and explored the inflammatory and transcriptomic correlates of higher concentrations.

## Methods

### Study Design and Participants

This cross-sectional study was approved by the Institutional Review Boards of The University of Texas Health Science Center at Houston and Memorial Hermann Hospital–Texas Medical Center (HSC-MS-25-0343) and was registered at ClinicalTrials.gov (NCT07157839). The cross-sectional study is reported in accordance with the Strengthening the Reporting of Observational Studies in Epidemiology (STROBE) reporting guideline.

The requirement for informed consent was waived for deidentified blood-bank specimens. Residual plasma specimens from 257 blood donors and fresh whole-blood specimens from 40 donors were collected at Memorial Hermann Hospital–Texas Medical Center between September 2025 and June 2026. In addition, 30 plasma samples from population reference participants and 20 from Alzheimer disease (AD) patients were purchased from BioIVT (Westbury, NY) and used to establish study-specific biomarker thresholds. Plasma samples from 738 participants in the University of Pittsburgh Alzheimer’s Disease Research Center, approved by the University of Pittsburgh IRB (STUDY 19110245), were used to evaluate the association between p-tau217 and age.

### Biomarker and RNA Measurements

Plasma p-tau217 was measured using nanoneedle assays with ALZpath and ADx antibodies; BD-tau, TNF-α, and IL-6 were measured using analyte-specific nanoneedle assays. Plasma specimens were analyzed in triplicate. Bulk RNA sequencing was performed on red blood cell–depleted fresh whole blood. Assay procedures, replicate-level quality control, handling of values outside assay limits, and bioinformatic methods are described in the *eMethods*.

### Statistical Analysis

Receiver operating characteristic (ROC) analyses of reference-population and AD samples were used to derive Youden thresholds for p-tau217 and BD-tau. The primary outcome was the proportion of donor plasma samples exceeding the p-tau217 Youden threshold. Note that the study-specific Youden thresholds were used as exploratory classification criteria rather than validated clinical reference limits. The secondary outcome was the proportion exceeding the prespecified published reference threshold for p-tau217. The Youden threshold was used for BD-tau, whereas published reference thresholds were used for TNF-α and IL-6. Biomarker associations were assessed using Kendall’s τ because distributions were skewed and below-LoD substitution resulted in tied values. Proportions were reported with Wilson 95% CIs. Spearman correlation was used for p-tau217–gene expression analyses and for the association between p-tau217 and age. Sensitivity analyses repeated biomarker correlations after excluding samples affected by assay failure, technical outliers, or values outside the quantifiable range. All tests were two-sided, with *P* < .05 considered statistically significant. Please see *eMethods* for the details.

## Results

### Plasma p-tau217 and BD-tau concentrations in population reference participants and AD patients

The population reference participants ranged in age from 18 to 59 years old (mean age 36 years), and half were women. The cohort comprised 13 Hispanic (43%), 13 African American (43%), and 4 non-Hispanic white (13%) participants. Body mass index (BMI) ranged from 18 to 64 kg/m² (mean: 28 kg/m²). Seven participants (23%) were current smokers, all of whom were male, while the remaining 23 (77%) were non-smokers. Blood types included O (16, 53%), A (6, 20.0%), B (7, 23%), and AB (1, 3%). Blood samples were collected between April and September 2024. The AD cohort consisted of 20 participants with a mean age of 77.6 years (range, 58–89 years); 12 (60%) were male and 8 (40%) were female. Participants included 10 Caucasian (50%), 8 African American (40%), and 2 Hispanic (10%) individuals. The mean MMSE score was 17.6 ± 2.5, and the mean ADAS-Cog score was 41.4 ± 14.0, indicating moderate cognitive impairment. Clinical Dementia Rating (CDR) scores were 1 in 10 participants (50%), 2 in 9 participants (45%), and 3 in 1 participant (5%), reflecting predominantly mild-to-moderate dementia severity. The 738 plasma samples were from participants enrolled in the PITT ADRC including 64.6% female; 83.2% non-Hispanic White; median age of 68.4 years [IQR 62.47-73.57]; median education 16 year [IQR: 12.00 to 18.00]).

Plasma p-tau217 and BD-tau concentrations were higher in AD patients than in population reference participants (**eFig. 1**). The study-specific Youden threshold was 1.24 pg/mL for p-tau217 measured by the nanoneedle using the ALZpath antibody, with 65% sensitivity and 87% specificity. For BD-tau, the ROC-derived Youden threshold was 39.85 pg/mL, with 90% sensitivity and 97% specificity (**eFig. 2**). For p-tau217, the range of prespecified published reference threshold published in major journals was 0.15 pg/mL -0.63 pg/mL ^1,2,16^.

### Plasma biomarker concentrations and threshold exceedance in blood donors

Using the study-specific ROC-derived Youden thresholds (**eFig. 2**), 4 of 30 reference samples (13%; 95% CI, 5%-30%) and 13 of 20 Alzheimer disease (AD) samples (65%; 95% CI, 43%-82%) exceeded the ALZpath Youden threshold. Corresponding proportions exceeding the published reference threshold (0.63 pg/mL ^2^) were 16 of 30 (53%; 95% CI, 36%-70%) for reference samples and 18 of 20 (90%; 95% CI, 70%-97%) for AD samples (**Fig. 1A**). For BD-tau, 1 of 30 reference samples (3%; 95% CI, 1%-17%) and 18 of 20 AD samples (90%; 95% CI, 70%-97%) exceeded the Youden threshold (**Fig. 1B**).

**Fig. 1.**
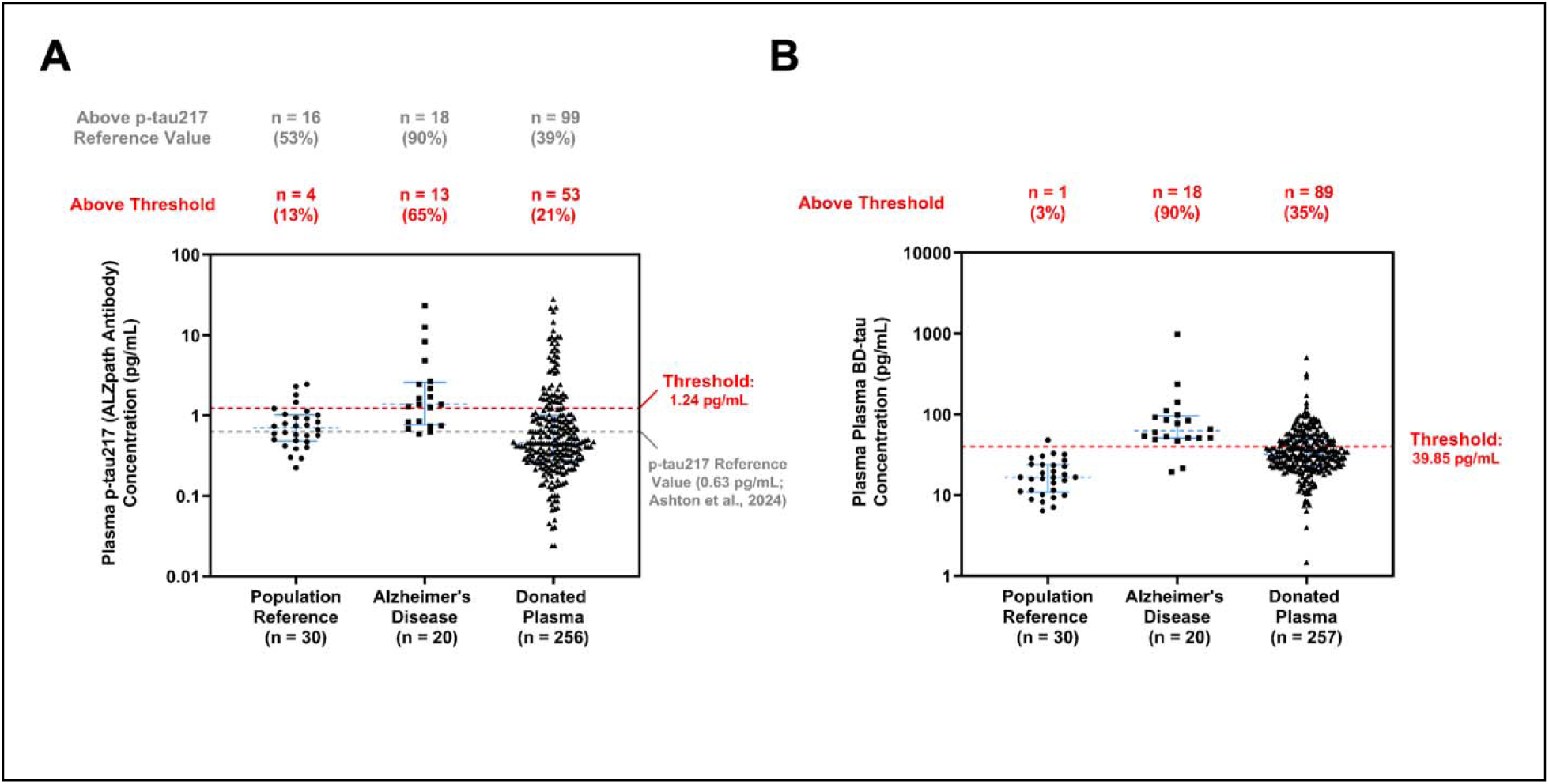
Plasma p-tau217 and BD-tau concentrations and exploratory threshold comparisons in blood donors, population reference participants, and Alzheimer’s disease patients. **A.** Plasma p-tau217 concentrations in blood donors were measured using nanoneedle assay with the ALZpath antibody (ALZpath Inc., Carlsbad, CA). The red dashed line represents the study-specific ROC-derived Youden threshold used for the primary outcome and the gray dashed line represents the prespecified published reference threshold ^2^ used for the secondary outcome. The number of blood donors with plasma p-tau217 concentrations above and below threshold values is shown. **B**. Plasma brain-derived tau (BD-tau) concentrations in blood donors were measured using nanoneedle assay with BD-tau antibody (Nanomosaic Inc., Waltham, MA). Red dashed line indicates the threshold value (the Youden threshold) established using population reference participants and Alzheimer’s disease patients. The numbers of blood donors with plasma BD-tau concentrations above and below the threshold value are shown. No between-group hypothesis tests were performed. p-tau217: tau protein phosphorylated at threonine 217; BD-tau: brain-derived tau.

**Table 1.** Demographic and Clinical Characteristics of the Population Reference Participants and Alzheimer’s Disease Cohorts.

| Characteristic | Population Reference (n = 30) | Alzheimer's Disease (n = 20) |
| --- | --- | --- |
| Age, years | 36 (18–59) | 78 (58–89) |
| Sex |  |  |
| Male | 15 (50%) | 12 (60%) |
| Female | 15 (50%) | 8 (40%) |
| Race/Ethnicity |  |  |
| African American | 13 (43%) | 8 (40%) |
| White | 4 (13%) | 10 (50%) |
| Hispanic | 13 (43%) | 2 (10%) |
| BMI, kg/m<sup>2</sup> | 28.3 (17.9–63.7) | — |
| Smoking status |  | — |
| Current smoker | 7 (23%) | — |
| Non-smoker | 23 (77%) | — |
| Blood type |  | — |
| O+ | 13 (43%) | — |
| O– | 3 (10%) | — |
| A+ | 4 (13%) | — |
| A– | 2 (7%) | — |
| B+ | 7 (23%) | — |
| AB+ | 1 (3%) | — |
| MMSE score | — | 17.6 (14–21) |
| ADAS-Cog score | — | 41.4 (17–74) |
| Clinical Dementia Rating (CDR) |  |  |
| CDR = 1 | — | 10 (50%) |
| CDR = 2 | — | 9 (45%) |
| CDR = 3 | — | 1 (5%) |

Plasma samples from 30 population reference participants and 20 Alzheimer's disease patients were commercially obtained from BioIVT (Westbury, NY, USA). Individual-level demographic and donor information, including age, sex, race/ethnicity, collection date, blood type, body mass index, and smoking status, was provided by BioIVT. Continuous variables are presented as mean (range), and categorical variables are presented as n (%). BMI, smoking status, and blood type were available only for the population reference cohort. MMSE, ADAS-Cog, and CDR were available only for the AD cohort. ADAS-Cog, Alzheimer's Disease Assessment Scale–Cognitive Subscale; BMI, body mass index; CDR, Clinical Dementia Rating; MMSE, Mini-Mental State Examination.

Notably, plasma p-tau217 concentrations exceeded the study-specific ROC-derived Youden threshold for the nanoneedle assay using the ALZpath antibody in 53 of 256 blood donors (21%; 95% CI, 16%-26%) and the published reference threshold in 99 of 256 donors (39%; 95% CI, 33%-45%) (**Fig. 1A**).

Plasma BD-tau concentrations exceeded the study-specific Youden threshold in 89 of 257 blood donors (35%; 95% CI, 29%-41%) (**Fig. 1B**). There was substantial interindividual variability in plasma TNF-α and IL-6 concentrations (**Fig. 2**). Plasma TNF-α concentrations exceeded the selected literature-derived reference values (6.0 pg/mL ^17^) in 103 of 255 blood donors (40%; 95% CI, 35%-47%), and plasma IL-6 concentrations exceeded the selected literature-derived reference values (7.0 pg/mL ^18^) in 129 of 254 blood donors (51%; 95% CI, 45%-57%) (**Fig. 2**).

**Fig. 2.**
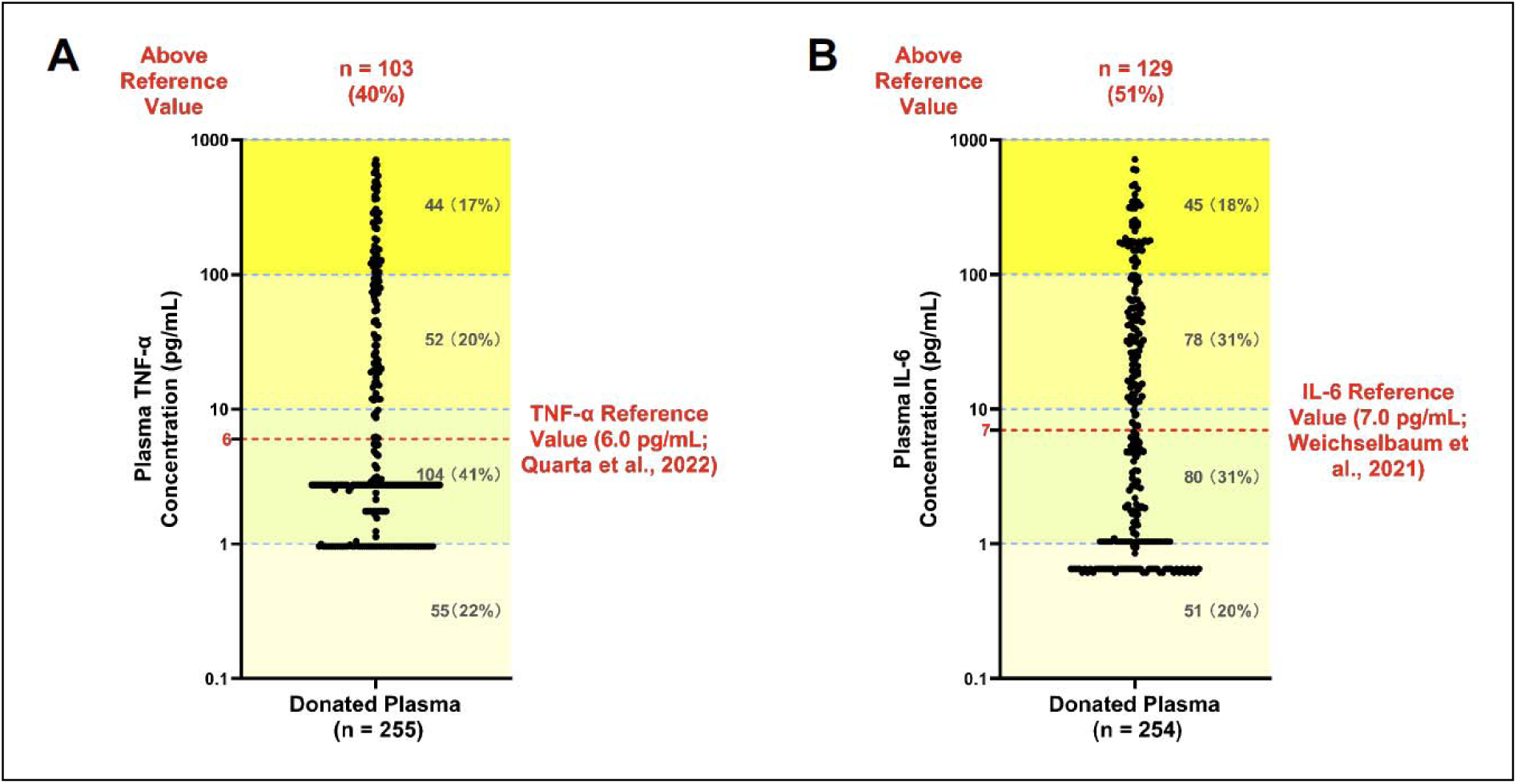
Plasma TNF-α and IL-6 concentrations and literature-derived reference comparisons in blood donors. **A**. Plasma tumor necrosis factor α (TNF-α) concentrations in blood donors. **B**. Plasma interleukin 6 (IL-6) concentrations in blood donors. The threshold value of TNF-α is derived from the study by Quarta et al ^17^. The threshold value of IL-6 is derived from the study by Weichselbaum et al ^18^. The numbers of blood donors with plasma TNF-α or IL-6 concentrations above and below the reference values are shown. No between-group hypothesis tests were performed. TNF-α: tumor necrosis factor α; IL-6: interleukin 6.

### Associations between plasma p-tau217 and inflammatory cytokines

Moreover, we found that the plasma p-tau217 concentrations were positively associated with TNF-α (**Fig. 3A**, τ = 0.339, *P* < .001, n = 254) and IL-6 (**Fig. 3B**, τ = 0.310, *P* < .001, n = 253) concentrations in blood donors. Interestingly, plasma BD-tau concentrations were only weakly positively associated with plasma p-tau217 concentrations (**eFig. 3**; τ = 0.132, *P* = 0.002, n = 256). Furthermore, plasma BD-tau concentrations demonstrated weakly positive associations with TNF-α (τ = 0.216, P < .001, n = 255) and IL-6 (τ = 0.183, *P* < .001, n = 254) concentrations (**eFig. 4**). Sensitivity analyses showed that the associations of p-tau217 (measured using the ALZpath antibody) with TNF-α and IL-6 remained significant after restricting the analysis to samples without assay failures, technical outliers, or values outside the quantifiable range (**eFig. 5**).

**Fig. 3.**
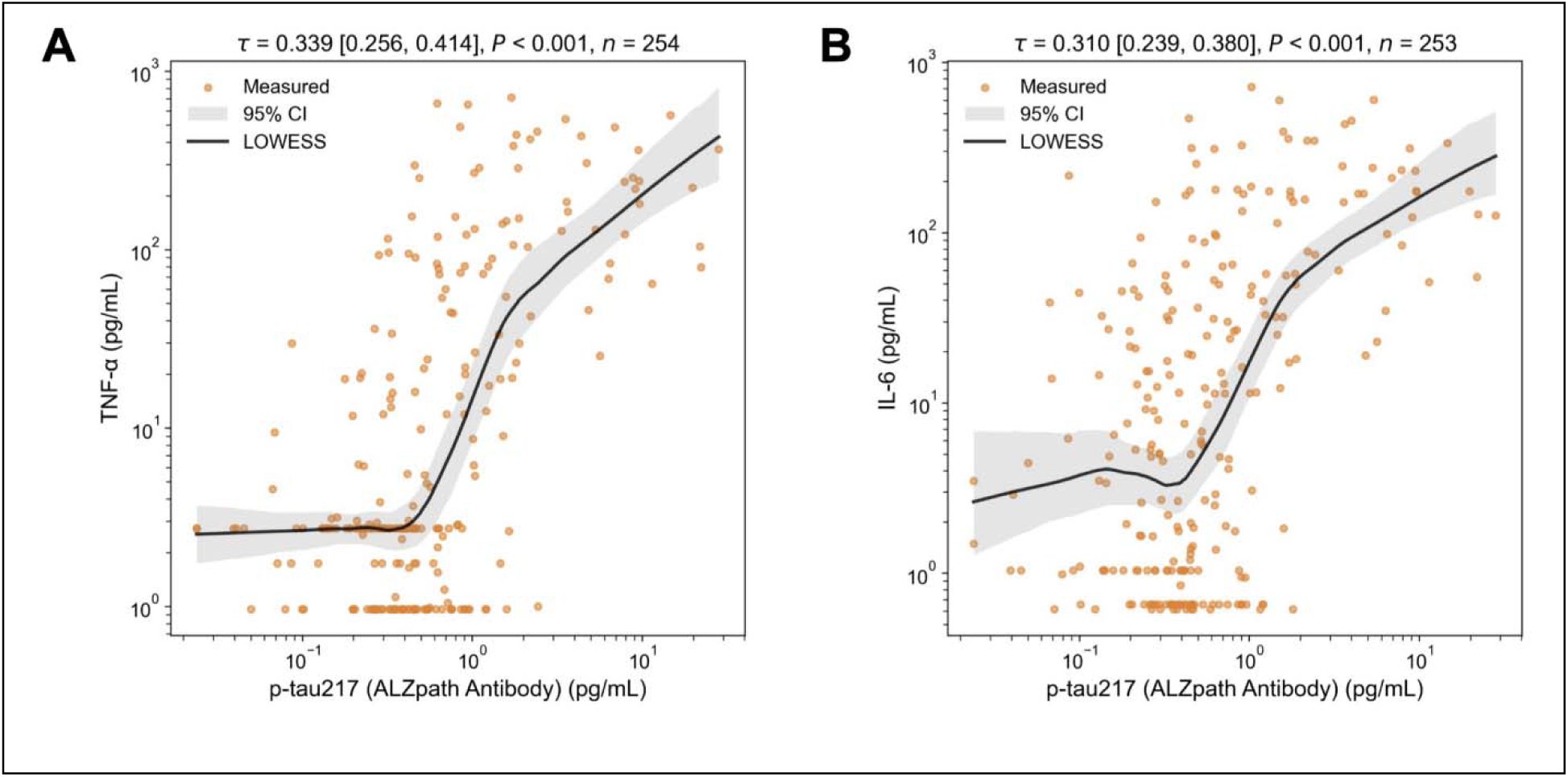
Associations between plasma p-tau217 and TNF-α or IL-6 in blood donors. **A**. Association between plasma p-tau217 (measured using the ALZpath antibody) and TNF-α concentrations in blood donors. **B**. Association between plasma p-tau217 (measured using the ALZpath antibody) and IL-6 concentrations in blood donors. Each point represents one donor sample. Solid black curves show locally weighted scatterplot smoothing (LOWESS) fits, and gray shaded bands indicate the corresponding 95% confidence intervals. LOWESS curves are provided to visualize trends; correlation coefficients and *P* values were calculated separately using Kendall’s τ. The τ values and *P* values represent the associations. p-tau217: Tau protein phosphorylated at threonine 217; TNF-α: tumor necrosis factor α; IL-6: interleukin 6.

### Measurement of p-tau217 using a different p-tau217 antibody

Next, we used a different antibody (ADx) to measure plasma p-tau217. ROC analyses using the ADx antibody identified a Youden threshold of 0.423 pg/mL. Using the ADx antibody, 164 of 253 blood donors (65%; 95% CI, 59%-70%) exceeded the study-specific ROC-derived Youden threshold of 0.423 pg/mL and 142 of 253 blood donors (56%; 95% CI, 50%-62%) exceeded the published reference threshold of 0.63 pg/mL ^2^ (**eFig. 6**).

Threshold exceedance was more frequent with ADx than ALZpath, indicating assay- and threshold-dependent classification. Applying the published ALZpath threshold (0.63 pg/mL ^2^) to ADx measurements was an exploratory cross-assay comparison.

Plasma p-tau217 concentrations measured using the ADx antibody remained positively associated with plasma TNF-α and IL-6 concentrations (**eFig. 7**). In addition, plasma p-tau217 concentrations measured using the ADx antibody showed weakly positive associations with plasma BD-tau concentrations (**eFig. 8**). Sensitivity analyses showed that the associations of p-tau217 (measured using the ADx antibody) with TNF-α and IL-6 remained significant after restricting the analysis to samples without assay failures, technical outliers, or values outside the quantifiable range (**eFigure 9**).

### Association between plasma p-tau217 concentrations and transcriptomic alterations

To explore transcriptomic correlates of higher plasma p-tau217 concentrations, we performed bulk RNA sequencing on fresh whole blood samples from 29 donors (**eFig. 10)** who had a wide range of plasma p-tau217 concentrations (**Fig. 4A**).

**Fig. 4.**
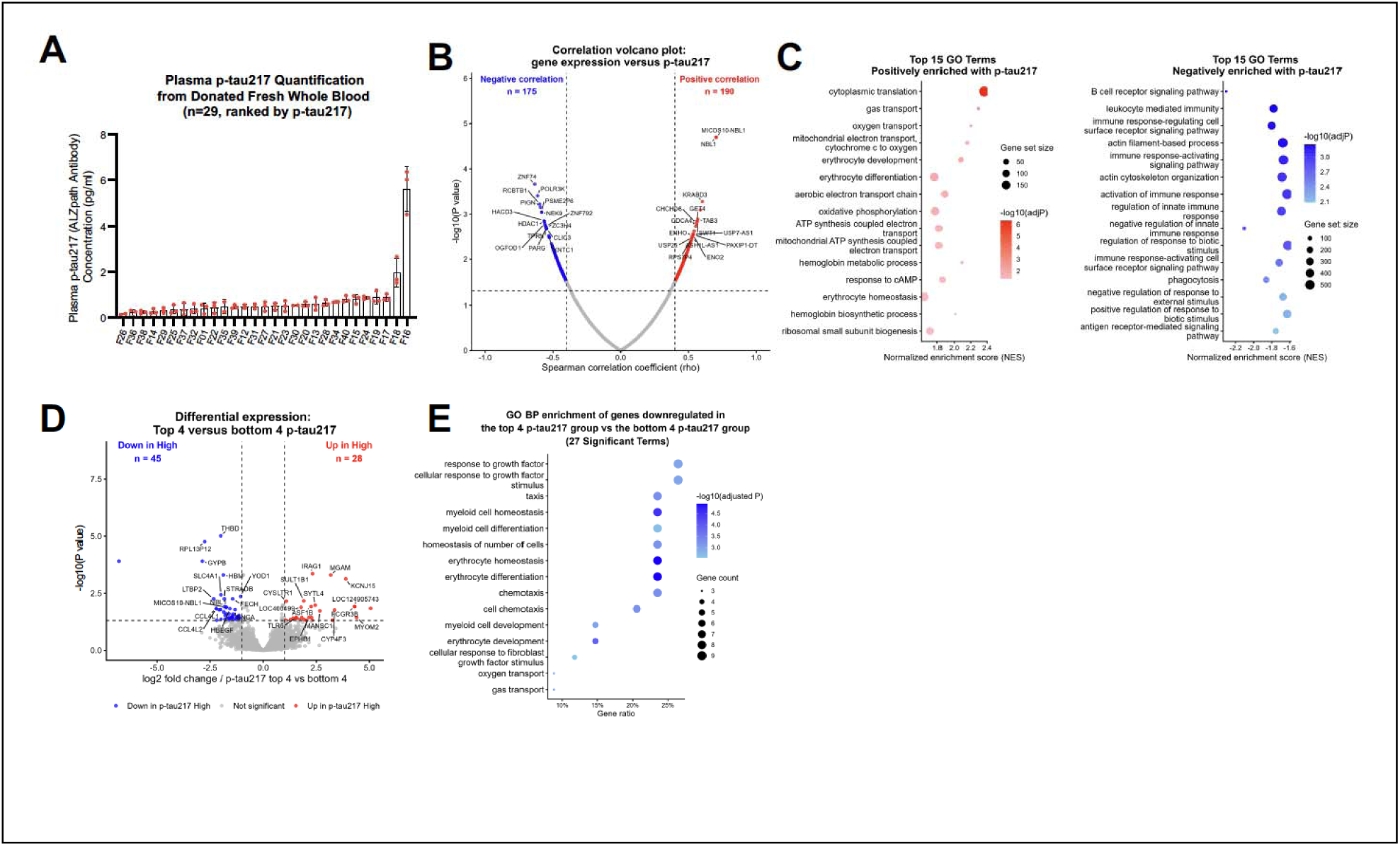
Bulk RNA sequencing in fresh whole blood of blood donors. **A.** The distribution of p-tau217 in the plasma of fresh whole blood samples obtained from 29 blood donors using ALZpath antibody to detect p-tau217. Error bars indicate standard deviation. **B**. Correlation volcano plot showing associations between fresh whole blood gene expression and plasma p-tau217 levels. Spearman correlation analysis was performed between variance-stabilized gene expression values and plasma p-tau217 concentrations across 29 samples. Each point represents one gene. The x-axis indicates the Spearman correlation coefficient (ρ), and the y-axis shows −log10(nominal *P* value). A total of 365 genes met the exploratory criteria of |ρ| > 0.4 and nominal *P* < .05, including 190 positively associated genes (red) and 175 negatively associated genes (blue). Gray points indicate genes that did not meet these criteria. The 15 genes with the smallest nominal *P* values in each direction are labeled. Dashed vertical lines indicate ρ = ±0.4, and the dashed horizontal line indicates nominal *P* = .05. Gene-level *P* values were not adjusted for multiple testing. **C**. Gene set enrichment analysis of GO Biological Process terms associated with p-tau217 levels. Genes were ranked according to their Spearman correlation coefficients with p-tau217, and GSEA was performed using Gene Ontology Biological Process gene sets. The dot plots show the 15 most significant positively enriched terms (left) and the 15 most significant negatively enriched terms (right), ranked by adjusted *P* value. Dot size represents the gene set size, and dot color represents −log10(adjusted *P*), with more intense colors indicating greater statistical significance. **D**. Fresh whole blood gene expression was compared between the top four and bottom four plasma p-tau217 levels. Differential expression analysis was performed using DESeq2. Genes with an absolute log2 fold change greater than 1 and an adjusted *P* value below 0.05 were considered differentially expressed. Red points indicate genes upregulated and blue points indicate genes downregulated in the top four p-tau217 samples compared to the bottom four p-tau217 samples, and gray points indicate genes that did not meet the significance thresholds. A total of 28 genes were upregulated and 45 genes were downregulated in the top 4 p-tau217 samples compared to the bottom 4 p-tau217 samples. The top 15 genes in each direction, ranked by *P* value, are labeled. Dashed vertical lines indicate log2 fold change = ±1, and the dashed horizontal line indicates an adjusted *P* value of 0.05. **E**. GO Biological Process enrichment analysis of genes downregulated in the top four p-tau217 group compared to bottom four p-tau217 group. GO Biological Process enrichment analysis was performed on genes downregulated in the top four p-tau217 samples compared with the bottom four p-tau217 samples. No significantly enriched GO terms were identified among the upregulated genes. A total of 27 significantly enriched terms were identified, and the 15 most significant terms ranked by adjusted *P* value are shown. Dot size represents the number of differentially expressed genes associated with each term, and dot color represents −log10(adjusted *P* value), with darker colors indicating greater statistical significance. GO: Gene Ontology; GSEA: Gene Set Enrichment Analysis; DESeq2: Differential Expression analysis for Sequence count data.

In an exploratory analysis, expression levels of 365 genes met the correlation criteria of |ρ| > 0.4 and nominal *P* < .05 for associations with plasma p-tau217 concentrations, including 190 positively associated and 175 negatively associated genes (**Fig. 4B**). These gene-level *P* values were not adjusted for multiple testing. Gene set enrichment analysis (GSEA) further identified multiple Gene Ontology (GO) Biological Process pathways associated with plasma p-tau217 levels (**Fig. 4C**).

To further characterize these transcriptional changes, we performed an exploratory extreme-phenotype analysis by comparing whole blood gene expression between participants with the top four and bottom four plasma p-tau217 levels. Differential expression analysis identified 73 differentially expressed genes, including 28 upregulated and 45 downregulated genes in the top four p-tau217 group compared to bottom four p-tau217 group (absolute log2 fold change > 1, adjusted *P* < .05; **Fig. 4D**). GO enrichment analysis identified 27 significantly enriched biological processes among the downregulated genes, whereas no enriched GO terms were detected among the upregulated genes (**Fig. 4E**). Measurement of p-tau217 using a different p-tau217 antibody (ADx) also yielded transcriptional changes, providing an additional exploratory analysis of the same RNA-sequencing dataset (**eFig. 11**).

Finally, plasma p-tau217 concentrations showed substantial interindividual variability across the adult age spectrum, with elevated concentrations observed in some participants beginning at approximately 30 years of age and across older age groups (**eFig. 12**).

## Discussion

In this exploratory cross-sectional study, plasma p-tau217 concentrations exceeded the study-specific ROC-derived Youden threshold in 21% of evaluable blood-donor samples. These findings raise the possibility that elevated circulating p-tau217 may occur in some individuals who meet blood-donation eligibility criteria. However, the threshold was derived from a small comparison cohort and has not been independently validated; therefore, these results do not establish the prevalence of clinically abnormal p-tau217 concentrations among blood donors.

Concentrations exceeding the respective study-specific thresholds were observed with both p-tau217 antibodies (**Fig. 1** and **eFig. 6**), although the proportions differed substantially. Positive associations with TNF-α and IL-6 were observed with both measurements (**Fig. 2** and **3**, **eFig. 5, 7** and **9)**. Furthermore, bulk RNA sequencing analyses of fresh whole blood from blood donors showed that plasma p-tau217 concentrations were associated with broad transcriptomic alterations involving multiple cellular pathways (**Fig. 4** and **eFig. 11**).

Elevated blood p-tau217 concentrations have been reported in a variety of conditions beyond AD, including cardiac arrest ^19^, Creutzfeldt–Jakob disease ^7^, chronic kidney disease ^9,10^, and anesthesia/surgery ^15^. Moreover, Gonzalez-Ortiz et al. reported the elevated plasma p-tau217 in newborns ^20^. However, the biological importance and potential consequences of elevations in plasma p-tau217 remain largely unexplored.

Higher plasma p-tau217 concentrations were associated with higher TNF-α and IL-6 concentrations and with exploratory patterns of blood-cell gene expression (**Fig. 3**). Expression levels of 365 genes met nominal correlation criteria (**Fig. 4**); however, these gene-level associations were not adjusted for multiple testing and require confirmation in an independent cohort.

Although a substantial proportion of blood donors exhibited concentrations exceeding the study-specific threshold, BD-tau showed only weak associations with TNF-α and IL-6 (**eFig. 4**). In contrast, p-tau217 demonstrated stronger relationships with these inflammatory markers.

Furthermore, the correlation between p-tau217 and BD-tau was weak (**eFig. 3** and **8**). Together, these findings suggest that plasma p-tau217 and BD-tau may capture partly distinct biological processes, with p-tau217 showing stronger associations with systemic inflammatory markers in this cohort.

The fresh whole blood bulk RNA sequencing studies (**Fig. 4** and **eFig. 11**) provide additional biological context. Although these data do not establish that circulating p-tau217 directly causes such cellular changes, they indicate that elevated circulating p-tau217 concentrations are associated with broad systemic transcriptomic alterations, although the direction and mechanisms of these associations remain unclear.

Intraoperative blood transfusions are associated with an increased risk of postoperative delirium in patients, although causality remains uncertain ^21^. Previous studies have raised concern about the potential transmission of amyloid-β pathology through blood transfusion ^22^. Epidemiologic data have shown an increased risk of intracerebral hemorrhage among recipients of red blood cells from donors who subsequently developed multiple intracerebral hemorrhages, a phenotype associated with cerebral amyloid angiopathy, although these observational findings do not establish causality ^23^. Recent report of cerebral amyloid angiopathy occurring decades after red blood cell transfusions has further raised this possibility ^24^. These observations have prompted consideration of whether other proteins associated with neurodegenerative diseases, including p-tau, could warrant similar investigation ^22^.

Whether circulating p-tau217 in donated blood has biological effects in transfusion recipients remains unknown. The present study did not evaluate transfused components, recipient exposure, or clinical outcomes and therefore cannot establish pathogenicity, transmissibility, or transfusion-related risk. The current blood transfusion practices remain safe and effective. Further work to characterize the molecular forms and biological activity of circulating p-tau217 could help determine whether recipient-focused studies are warranted.

As a contextual analysis, we examined plasma p-tau217 concentrations across the adult age spectrum in an independent PITT-ADRC cohort. Substantial interindividual variability in plasma p-tau217 concentrations was observed across age groups, including among some young adults (**eFig. 12**). However, this analysis was not intended to validate the prevalence estimates observed in blood donors because donor demographic characteristics were unavailable, and the PITT-ADRC and blood-donor samples differed in participant characteristics, sample collection and processing procedures, and p-tau217 assay platforms. Thus, the PITT-ADRC findings only provide contextual evidence that plasma p-tau217 concentrations vary substantially across adulthood.

The blood donor samples were obtained from a single academic medical center blood bank in the United States and their characteristics were unavailable due to de-identification. However, about 63% of blood donors are between 25 and 64 years old, and 16% are more than 65 years old ^25^. The extent to which observed elevations in plasma p-tau217 and BD-tau are generalizable remains unknown. Additionally, blood collection, processing, storage, and donor-screening practices may vary among blood banks and blood collection organizations, potentially influencing biomarker measurements. Blood-donation eligibility does not establish the absence of neurological or systemic conditions that could influence biomarker concentrations, and the generalizability of these findings remains uncertain. It remains unknown whether apparent elevations in circulating p-tau217 and BD-tau are normal, prognostic, or diagnostic for sub-clinical disease.

The relatively small population reference cohort also limited our ability to establish robust population-based reference ranges. In addition, amyloid positivity may provide a more biologically relevant classification of elevated p-tau217; however, amyloid status was not available for participants assessed with the nanoneedle assays. Future studies incorporating amyloid biomarkers and harmonized assay platforms in larger, well-characterized populations are needed to establish and validate clinically and biologically meaningful thresholds.

Plasma p-tau217 thresholds vary among assays because of differences in antibodies, platforms, and calibration, ranging from 0.15 to 0.63 pg/mL ^1,2,16,26^. The published threshold was used only for an exploratory secondary comparison. Its applicability to the nanoneedle assay has not been established; therefore, exceedance should not be interpreted as evidence of amyloid positivity or clinically abnormal p-tau217 concentrations.

A substantial proportion of TNF-α and IL-6 measurements were below the limit of detection (LoD; **eTable 1**). Nevertheless, their associations with p-tau217 remained statistically significant after excluding samples with below-LoD measurements (**eFigs. 5** and **9**). The difference in donor threshold exceedance between ALZpath (21%) and ADx (65%) reflected both assay-dependent concentration measurements and different threshold operating points, with respective specificities of 87% and 67% (**eTable 2**). ADx generally yielded higher concentrations, and differences persisted when the same numerical cutoff was applied. Plasma p-tau217 concentrations measured using the ALZpath and ADx antibodies were positively correlated (**eFig. 13**).

A limitation is that our cross-sectional design precludes assessing consequences of elevated plasma p-tau217 and BD-tau in blood donors. The population reference participants and blood donors differed in terms of participant characteristics. For example, the population reference participants were not confirmed to be AD-negative and therefore may have included individuals with preclinical AD and elevated p-tau217 levels. Additionally, the population reference participants were substantially younger than the participants with AD (mean age, 36 vs. 78 years). Comparison between AD and non-AD groups was not age-matched, thus, the observed biomarker differences between the AD and reference groups may partly reflect their substantial age difference. Finally, the mechanistic links among circulating p-tau217, BD-tau, inflammation, and transcriptomic alterations remain to be determined.

In conclusion, in this exploratory study, plasma p-tau217 and BD-tau concentrations exceeded study-specific thresholds in a subset of blood-donor samples. Higher p-tau217 concentrations were associated with inflammatory biomarkers and exploratory transcriptomic differences. These observations raise the possibility of elevated circulating tau species in some blood donors and warrant confirmation in well-characterized cohorts using validated assay-specific thresholds.

## Data Availability

All data produced in the present study are available upon reasonable request to the authors

## Acknowledgments

These studies were performed at The University of Texas Health Science Center at Houston, and are attributed to the Department of Anesthesiology, Critical Care and Pain Medicine, Institute of Perioperative Medicine, McGovern Medical School, The University of Texas Health Science Center at Houston.

## Declaration of Generative AI Use

During the revision of this manuscript, the authors used ChatGPT (OpenAI) to improve the clarity, grammar, and readability of the text. The authors reviewed and edited all AI-assisted content and take full responsibility for the final manuscript.

## Data Sharing Statement

Deidentified blood-donor biomarker data, the corresponding data dictionary, processed RNA-sequencing gene-count matrices, and analytical code supporting the findings will be available upon publication of the manuscript. Requests should be directed to Zhongcong Xie, MD, PhD, at and should include a description of the proposed research and requested materials. Access will be provided for scientifically sound proposals, subject to applicable institutional requirements and data-use agreements. Access to data obtained from the University of Pittsburgh Alzheimer’s Disease Research Center and commercial sample providers will be governed by the respective providers’ sharing policies and agreements.

## Funding Sources

This research was partially supported in design and performance of studies, data analysis and manuscript preparation by R01AG098122 to Zhongcong Xie, R35GM166431 to Yiying Zhang, R01HL165748, R01HL169519, R35HL177402, T32GM135118, and R24HL180372 to Holger K.

Eltzschig from the National Institutes of Health, Bethesda, MD. The National Institutes of Health had no role in the design and conduct of the study; collection, management, analysis, and interpretation of the data; preparation, review, or approval of the manuscript; or decision to submit the manuscript for publication.

## Author contributions

ZX, XL, WZ, RW, HT, SF and QQ: Conceived and designed the project.

XL, WZ, RW, HT, SF and CT: Performed the experiments, analyzed the data, and prepared the figures.

ZX, XL and WZ wrote the manuscript.

ZX, DIS and TK revised the manuscript.

FL, XY, YL, YJ, WC, YD, YSZ, SS, YZ, LM, HKE, and AT provided critical comments on the study design, data analysis and revision of the manuscript.

All authors reviewed the manuscript before the submission of the manuscript.

Dr. Zhongcong Xie had full access to all the data in the study and takes responsibility for the integrity of the data and the accuracy of the data analysis.

