## Supplemental methods and figures for "Elevation in Plasma p-tau217 Concentrations Among Blood Donors: An Exploratory Cross-Sectional Study"

Pittsburgh, PA 15213, USA

Current Address of Xiang Li: Department of Anesthesia, Shanghai General Hospital, Shanghai, P.R. China.
Current Address of Feng Liang: Center for Medical-Engineering Cross-Disciplinary Research, the Institute of Technological Science, Wuhan University, Wuhan, China.

Current Address of Yuanlin Dong: Department of Neurology, Brigham and Women's Hospital and Harvard Medical School, Boston, MA 02124

### Index of Supplemental Materials

**eMethods.**

**eFig. 1.** Plasma p-tau217 and BD-tau concentrations in reference participants and patients with Alzheimer's disease.

**eFig. 2.** Derivation of study-specific plasma p-tau217 and BD-tau thresholds.

**eFig. 3.** Association between plasma p-tau217 and BD-tau concentrations in blood donors.

**eFig. 4.** Associations between plasma BD-tau and TNF-α or IL-6 in blood donors.

**eFig. 5.** Sensitivity analyses of associations between plasma p-tau217 and inflammatory biomarkers.

**eFig. 6.** Plasma p-tau217 concentrations and threshold comparisons using the ADx antibody.

**eFig. 7.** Associations between plasma p-tau217 and TNF-α or IL-6 in blood donors using a different antibody (ADx).

**eFig. 8.** Association between plasma p-tau217 and BD-tau using a different antibody (ADx).

**eFig. 9.** Sensitivity analyses of associations between plasma p-tau217 and inflammatory biomarkers.

**eFig. 10.** Flowchart of fresh whole blood sample processing for plasma p-tau217 measurement and bulk RNA sequencing.

**eFig. 11.** Exploratory associations between blood-cell gene expression and plasma p-tau217 measured using the ADx antibody.

**eFig. 12.** Distribution of plasma p-tau217 concentrations across age in cognitively normal individuals.

**eFig. 13.** Association between plasma p-tau217 measured by ALZpath and ADx antibodies.

**eTable 1.** Summary of flagged replicate measurements.

**eTable 2.** Comparison of p-tau217 measurements and threshold exceedance between ALZpath and ADx antibodies.

**eMethods**

**Participants**

The participants were individual blood donors. De-identified blood segments were collected from donated units at the blood bank of Memorial Hermann Hospital–Texas Medical Center in Houston, Texas. As donor identities and characteristics were unavailable due to Health Insurance Portability and Accountability Act of 1996 (HIPAA) restrictions, no additional research-specific inclusion or exclusion criteria were applied beyond the standard eligibility requirements for blood donation. The institutional review board waived the requirement for informed consent because the study used deidentified samples and involved minimal risk to participants.

We purchased 30 plasma samples of population reference participants and 20 plasma samples of AD patients from BioIVT (Westbury, NY). Plasma samples were obtained from 257 blood donors and fresh whole blood samples were obtained from 40 blood donors at the blood bank of Memorial Herman Hospital, Houston, Texas. Donor identities and characteristics were not available due to HIPAA restrictions. Additional 738 plasma samples from participants enrolled in the University of Pittsburgh Alzheimer's Disease Research Center (PITT-ADRC) were used to evaluate the relationship between p-tau217 and age. Details of the PITT-ADRC study have been described previously [1]. The study was approved by the University of Pittsburgh Institutional Review Board (STUDY 19110245), and all participants provided written informed consent.

**Blood Samples**

Plasma and fresh whole blood samples were obtained from the Memorial Hermann Hospital Blood Bank between September 2025 and June 2026, based on sample availability and research personnel availability for specimen collection and processing. The blood center prepared small segments from plasma and fresh whole-blood products for blood typing. Each segment was detached from the tubing by twisting. Plasma segments were stored at 1–6°C at the Memorial Hermann Hospital–Texas Medical Center Blood Bank and transported to the testing laboratory at McGovern Medical School at 1–6°C. Citrate-based anticoagulant solutions, including citrate-phosphate-dextrose-adenine (CPDA-1), citrate-phosphate-dextrose (CPD), or acid-citrate-dextrose (ACD), were used as the primary anticoagulants. Upon arrival at the laboratory, the samples were immediately centrifuged at 12,000 × *g* for 20 minutes. The resulting supernatant was collected and stored at −80°C until enough samples had accumulated for batch analysis. The interval from sample storage to measurement was from 1 to 4 weeks.

Fresh whole blood remaining in tubing segments used for blood typing was collected and processed immediately upon arrival by centrifugation to separate plasma and cellular components. To separate components of fresh whole blood, we first performed extraction of total plasma and white blood cells. Blood was centrifuged at 1000 g for 10 minutes to obtain the plasma supernatant. The resulting pellet was then treated with a 1:9 ACK solution to remove red blood cells at room temperature for 5 minutes, followed by centrifugation to discard the supernatant. The subsequent pellet was then treated again with a 1:9 ACK solution at room temperature for 5 minutes, and the final pellet was collected as the white blood cells. Total RNA was then extracted from the cells using TRIzol reagent and RNeasy Universal Kit, and the purified RNA was sent for RNA sequencing. The plasma samples were stored at −80°C until analysis.

Purchased plasma samples of AD and population reference cohorts were used only to establish reference thresholds, whereas the primary study population consisted of blood donors in the present study. The plasma samples from participants enrolled in PITT-ADRC were used to determine the distribution of p-tau217 concentrations across the adult age spectrum.

**Variables**

The primary variables were plasma phosphorylated tau at threonine 217 (p-tau217), brain-derived tau (BD-tau), tumor necrosis factor α (TNF-α), and interleukin 6 (IL-6) concentrations. Plasma p-tau217 was measured using a nanoneedle assay with the ALZpath p-tau217 antibody (ALZpath Inc., Carlsbad, CA, USA), with independent measurement using the ADx p-tau217 antibody (ADx NeuroSciences, Ghent, Belgium). Plasma BD-tau concentrations were measured using a nanoneedle assay with a BD-tau–specific antibody (NanoMosaic, Waltham, MA) [2,3]. Plasma TNF-α and IL-6 concentrations were measured using nanoneedle assays with anti-human TNF-α (Catalog #DY210) and anti-human IL-6 (Catalog #DY206) antibodies obtained from R&D Systems (Minneapolis, MN). TNF-α and IL-6 served as biomarkers of systemic inflammation. The cellular components of fresh whole blood were used to extract RNA, which was used for bulk RNA sequencing. Biomarker concentrations were analyzed as continuous variables, and elevated biomarker concentrations were defined by predefined thresholds.

For p-tau217, elevated concentrations were operationally defined as values exceeding the study-specific ROC-derived Youden threshold. This definition was used to describe the sampled blood donor specimens and does not establish clinical abnormality, amyloid positivity, or Alzheimer's disease. The published threshold was examined as an exploratory secondary comparison because its applicability to the nanoneedle assay has not been established. For BD-tau, the Youden index threshold was established using ROC analysis of population reference participants and AD patient samples. For TNF-α [4] and IL-6 [5], elevated concentrations were defined per published values.

**Biomarker and RNA Measurements**

Plasma p-tau217 was measured using a nanoneedle assay with the ALZpath antibody (ALZpath Inc., Carlsbad, CA, USA), with independent determination using the ADx antibody (ADx NeuroSciences, Ghent, Belgium). BD-tau was measured using a nanoneedle assay with a BD-tau–specific antibody (NanoMosaic, Waltham, MA), whereas TNF-α (#DY210) and IL-6 (#DY206) were measured using nanoneedle assays with antibodies from R&D (Minneapolis, MN). Calibration standards were analyzed in duplicate and plasma samples in triplicate. Replicate-level quality control was performed before concentration calculation, and markedly discrepant technical replicates were excluded. Limit of detection (LoD) for each plate was derived from blank and low-concentration calibrator signals and converted to concentrations using the corresponding standard curves. Values below the LoD were assigned LoD for primary statistical analyses; values above the upper limit of quantification were excluded. RNA isolated from red blood cell–depleted fresh whole blood underwent bulk RNA sequencing, quality control, alignment to GRCh38, gene-level quantification, and DESeq2 processing.

**Study samples and assays**

We collected 257 plasma specimens and 40 fresh whole blood specimens from blood donors, 30 plasma specimens from the reference population participants, and 20 plasma specimens from AD patients. Prior to analysis, plasma aliquots were thawed and centrifuged at 14,000 × g for 5 minutes at 4°C. Baseline signals were recorded for each nanoneedle plate prior to antibody immobilization. Capture antibodies were then immobilized on the nanoneedle sensor surface overnight, followed by blocking with blocking buffer and incubation with plasma samples or calibration standards. Detection antibodies were subsequently applied, and signals were amplified and developed according to the manufacturer's nanoneedle assay protocol. After signal acquisition, net signals were calculated by subtracting baseline signals from post-development signals. Quantitative measurements were obtained using the Tessie nanoneedle platform, with calibration standards analyzed in duplicate and plasma samples analyzed in triplicate.

*Handling of Values Outside the Quantifiable Range*

The limit of detection (LoD) was determined on a plate-specific basis using the corresponding standard curve data for each assay plate. For each plate, the limit of blank (LoB) was first calculated using the signals from the blank wells:

$$LoB_{signal} = mean_{blank} + 1.645 \times SD_{blank}$$

where "$mean_{blank}$" represents the mean signal of the blank wells and "$SD_{blank}$" represents the standard deviation of the blank-well signals. The coefficient 1.645 corresponds to the one-sided 95th percentile of the standard normal distribution.

The LoD signal was then calculated as follows:

$$LoD_{signal} = LoB\_signal + 1.645 \times SD_{low}$$

where "$SD_{\mathrm{low}}$" represents the standard deviation of the signal values at the lowest non-zero calibrator concentration with acceptable precision. In this study, acceptable precision was defined as a coefficient of variation (%CV) ≤ 20%. If the %CV of the lowest non-zero calibrator exceeded 20%, the next higher non-zero calibrator concentration with a %CV≤20% was selected sequentially, and the standard deviation of its signal values was used as $SD_{\mathrm{low}}$.

The calculated ($LoD_{signal}$) was subsequently converted to a concentration using the corresponding plate-specific standard curve. Specifically, two adjacent calibrator concentrations whose mean signal values bracketed the calculated ($LoD_{signal}$) were identified, and the corresponding LoD concentration ($LoD_{concentration}$) was determined by linear interpolation of two adjacent calibrators. If ($LoD_{signal}$) fell between the mean signal of the blank wells and that of the lowest non-zero calibrator, interpolation was performed using the blank point and the lowest non-zero calibrator point.

For statistical analyses, sample concentrations falling below the LoD were assigned a value of LoD, calculated by $LoD_{concentration}/\sqrt{2}$, a substitution method established by Hornung and Reed [6] to minimize bias when estimating the true mean and variance of moderately skewed, log-normally distributed data. For the primary analysis, replicate concentrations below the plate-specific LoD were replaced with that LoD before calculating each sample's mean concentration. Sensitivity analyses excluded samples with any below-LoD replicate measurement for either analyte included in the correlation.

The upper limit of quantification (ULOQ) was defined as the highest calibrator concentration within the standard curve range that could be reliably quantified. Samples with signals exceeding the upper quantifiable range of the standard curve, or for which the software reported concentrations as too high to be reliably estimated, were considered above the ULOQ. These values were neither extrapolated beyond the standard curve nor replaced with the ULOQ. The original assay results were retained for quality-control records but were excluded from subsequent statistical analyses.

*Replicate-Level Quality Control*

Each analyte was measured in triplicate for each sample, with assay signals expressed in Nano Units. Measurements that failed instrument detection were reported as "not quantifiable" and were excluded. For replicate-level quality control, each replicate's normalized deviation was calculated by dividing the absolute difference between its signal and the median signal of the three technical replicates by that median. Replicates differing from the median by more than approximately 70.7% were classified as technical outliers and excluded from the analysis. The final concentrations were derived from the mean of non-outliers with the minimal number of valid replicates as one. Any samples below LoD were treated as LoD. Each sample's final concentration was calculated as the mean of its retained replicate concentrations, with at least one retained replicate required. Measurements above the upper limit of quantification (ULOQ) were excluded; the remaining valid replicate measurements with minimal number of one were used to calculate the mean concentration for each sample.

*Assay Precision Assessment*

For the p-tau217 assay using the ALZpath antibody, 257 donated human plasma samples were included, yielding a mean intra-CV of 15%. The calculated value was obtained from nano unit signals after exclusion of outlier. For the p-tau217 assay using the ADx antibody, 250 samples were included, resulting in a mean intra-CV of 13%. Seven samples were excluded because of assay failure: samples M085, M104, M112, M127, M170, and M230 each had two failed measurements among three technical replicates, and sample M210 had all three replicate measurements fail. For p-tau217 measurements in plasma prepared from fresh whole blood, the mean intra-CV was 15%, calculated from 38 samples because p-tau217 was below the limit of detection (LoD) in one sample (No. 16) and failure to harvest plasma from one sample (No. 6).

Three samples (Nos. 5, 10, and 21) had individual replicate p-tau217 measurements above the ULOQ. Only the above-ULOQ measurements were excluded; the remaining valid replicate measurements were used to calculate the mean concentration for each sample. For the BD-tau assay, all 257 donated human plasma samples were included, yielding a mean intra-CV of 20%. For the TNF-α assay, 256 samples were included, resulting in a mean intra-CV of 11%; sample M170 was excluded because all three technical replicate measurements failed. For the IL-6 assay, all 257 donated human plasma samples were included, yielding a mean intra-CV of 11%.

ROC curve analyses were performed using plasma samples from 30 population reference participants and 20 Alzheimer's disease (AD) patients to evaluate the ability of plasma p-tau217 and BD-tau to discriminate between the two groups. For each biomarker, the optimal threshold value was determined using the Youden index. The resulting threshold values were subsequently applied to the donated human plasma samples to determine the proportion of samples with biomarker concentrations above the corresponding threshold.

Associations between plasma biomarkers were assessed using Kendall's rank correlation coefficient (Kendall's $\tau$). Kendall's $\tau$ was selected because a substantial proportion of TNF-α and IL-6 concentrations were below LoD and were assigned the corresponding plate-specific LoD concentrations, resulting in tied values. Correlation coefficients are reported as Kendall's $\tau$ with 95% confidence intervals (CIs).

*Sensitivity Analysis*

Sensitivity analyses were performed using a restricted dataset containing only samples that met all predefined quality-control and quantification criteria. At the sample level, a sample was excluded from the sensitivity analysis if any measurement for the relevant analyte exhibited assay failure, was identified as a markedly discrepant technical replicate during replicate-level quality control, or yielded a concentration below the plate-specific LoD or above the ULOQ. Thus, unlike the primary analysis, in which valid replicate measurements were retained after replicate-level quality control and values below the LoD were replaced with the corresponding plate-specific LoD concentration, the sensitivity analysis excluded the entire affected sample.

Kendall's tau correlation analyses were then repeated using the restricted dataset. The resulting correlation coefficients, 95% CIs, and *P* values were compared with those from the primary analysis to assess the robustness of the observed associations to assay quality and the handling of values outside the quantifiable range.

Blood samples of PITT-ADRC participants were collected by venipuncture into EDTA tubes and processed to obtain plasma for biomarker measurements followed standardized procedures [7]. Plasma p-tau217 levels were measured using the ALZpath p-tau217 V2 assay (#104371) on the SIMOA HD-X platform (Quanterix, Billerica, MA). Prior to analysis, plasma samples were thawed and centrifuged at 4,000 × g for 10 minutes to remove particulates. Three quality control (QC) samples were run in duplicate at the beginning and end of each run to monitor assay performance. The mean within-run coefficient of variation was 8.1% versus 6.9% between-run. All samples were run in singlet.

**RNA-seq quality control and data processing**

Red blood cells were lysed before RNA extraction. Total RNA was extracted using a RNeasy Universal Kit (Qiagen, Germantown, MD, USA), and RNA sequencing was performed by Plasmidsaurus, Inc. (Louisville, KY, USA). Raw single-end sequencing reads were assessed using FastQC. Adapter trimming and quality filtering were performed using Trim Galore with a minimum Phred quality score of 20 and a minimum retained read length of 20 nucleotides. Read quality was reassessed using FastQC after trimming. The GRCh38 primary human genome assembly and GENCODE release 49 basic gene annotation were used for read alignment and gene-level quantification. Trimmed reads were aligned to the GRCh38 reference genome using STAR, and coordinate-sorted BAM files were generated.

Gene-level counts were generated using featureCounts v2.1.1 with the GENCODE v49 basic annotation. Samples with a uniquely mapped read rate below 70% were excluded. Count data from the remaining samples were imported into DESeq2, and sample-specific size factors were estimated using the median-of-ratios method. Samples with a size factor below 0.5 or above 2.0 were removed. Following sample-level quality control, low-expression genes were filtered out by retaining only genes with a raw count greater than 10 in at least half of the remaining samples. Size factors and dispersion parameters were re-estimated after filtering. After sample and gene filtering, variance-stabilizing transformation was performed using the DESeq2 varianceStabilizingTransformation function.

**Spearman correlation analysis and gene set enrichment analysis (GSEA)**

Spearman correlation analysis was performed to evaluate the associations of gene expression in red blood cell–depleted blood samples with plasma p-tau217 levels measured using the ALZpath antibody or ADx antibody. For each retained gene, Spearman correlation coefficients and corresponding two-sided *P* values were calculated using variance-stabilized expression values across the final samples (n = 29). Genes with an absolute Spearman correlation coefficient greater than 0.4 and a *P* value below 0.05 were considered correlated with p-tau217. Nominal *P* values were used for exploratory visualization of individual gene-level associations as described in previous studies [8-10].

For gene set enrichment analysis, all genes were ranked according to their Spearman correlation coefficients with plasma p-tau217 levels. GSEA was then performed using Gene Ontology Biological Process gene sets. Positive normalized enrichment scores indicated biological processes positively associated with p-tau217, whereas negative normalized enrichment scores indicated processes inversely associated with p-tau217. Enrichment *P* values were adjusted using the Benjamini–Hochberg method, and terms with an adjusted *P* value below 0.05 were considered statistically significant.

**Differential gene expression analysis between the top and bottom four p-tau217 groups and GO Biological Process enrichment analysis**

To examine gene-expression differences between individuals with extreme plasma p-tau217 levels, the four samples with the highest p-tau217 levels (top four) and the four samples with the lowest p-tau217 levels (bottom four) were selected from the quality-controlled cohort. Differential expression analysis was performed using DESeq2. Fold changes were calculated as the top four p-tau217 samples relative to the bottom four p-tau217 samples. *P* values were adjusted for multiple comparisons using the Benjamini–Hochberg method. Genes with an adjusted *P* value below 0.05 and an absolute log2 fold change greater than 1 were considered differentially expressed.

Gene Ontology Biological Process over-representation analysis was performed separately for significantly upregulated and significantly downregulated genes in the top four p-tau217 samples compared to the bottom four p-tau217 samples. Differentially expressed genes were defined as those with an adjusted *P* value below 0.05 and an absolute log2 fold change greater than 1. Enrichment *P* values were adjusted for multiple testing using the Benjamini–Hochberg method, and GO terms with an adjusted *P* value below 0.05 were considered significantly enriched.

**Sample size**

Sample size was determined by the availability of de-identified residual blood-bank specimens during the study period rather than by a formal power calculation. Because this was an exploratory cross-sectional study, the primary outcome was the proportion of donor samples exceeding the study-specific ROC-derived Youden threshold. The available sample of approximately 250 donor specimens provided 95% CIs with margins of error of approximately 5 to 6 percentage points for estimated proportions of sampled donor specimens exceeding the threshold in the range observed in this study.

**Statistical Analysis**

Continuous variables are presented as individual values with means ± standard deviations (SD). Categorical variables are presented as numbers and percentages. Comparisons between groups in eFig. 1 and eFig. 6 were performed using Mann-Whitney test.

For p-tau217, as a primary analysis, ROC curve analysis using population reference participants and AD patients was performed to derive a study-specific Youden threshold, which was subsequently applied to the blood donor samples. The prespecified published reference threshold was applied to determine the secondary outcome, defined as the proportion of blood donor samples exceeding this threshold.

Associations among plasma p-tau217, BD-tau, TNF-α, and IL-6 concentrations were evaluated using Kendall's $\tau$ correlation analysis because biomarker distributions were skewed and included values near or below the assay detection limits. Spearman correlation analysis was performed between the variance-stabilized expression values and the corresponding p-tau217 levels across fresh whole blood samples for each gene. The correlation between plasma p-tau217 concentration and age among PITT-ADRC participants was estimated using Spearman rank correlation. Proportions were reported with 95% CIs calculated using the Wilson score method. Correlation coefficients and two-sided $P$ values were reported.

Biomarker concentrations were analyzed as continuous variables; predefined thresholds were used to estimate the proportion of donor samples exceeding the selected thresholds. No adjustment for donor demographic or clinical covariates was performed because donor-level characteristics were unavailable.

Statistical analyses were performed using GraphPad Prism version 10 (GraphPad Software, San Diego, CA), R version 4.5.2 (R Foundation for Statistical Computing, Vienna), and Python version 3.13.9 (Python Software Foundation, Wilmington, DE). All statistical tests were two-sided, and $P < .05$ was considered statistically significant unless otherwise specified.

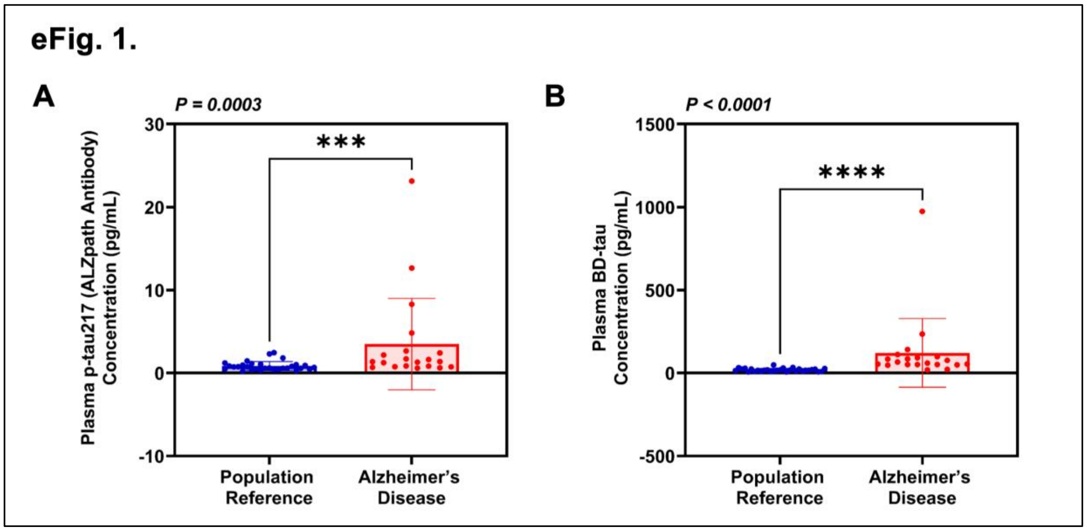


**eFig. 1. Plasma p-tau217 and BD-tau concentrations in reference participants and patients with Alzheimer's disease.**

**A**. Plasma p-tau217 concentrations in population reference participants and Alzheimer's disease patients were measured using nanoneedle assays with the ALZpath antibody (ALZpath Inc., Carlsbad, CA). **B**. Plasma BD-tau concentrations in population reference participants and patients with Alzheimer's disease were measured using nanoneedle assays with the BD-tau antibody (Nanomosaic Inc., Woburn, MA). The utilization of ALZpath antibody and BD-tau antibody demonstrated that plasma p-tau217 and BD-tau concentrations were higher in the purchased AD samples than in the reference samples. These comparisons describe group differences within this sample set. Mann–Whitney test was used to analyze the data. Error bars indicate standard deviation. The *P* values represent the differences in concentrations of p-tau217 and BD-tau between population reference participants and Alzheimer's disease patients. p-tau217: tau protein phosphorylated at threonine 217; BD-tau: brain-derived tau.

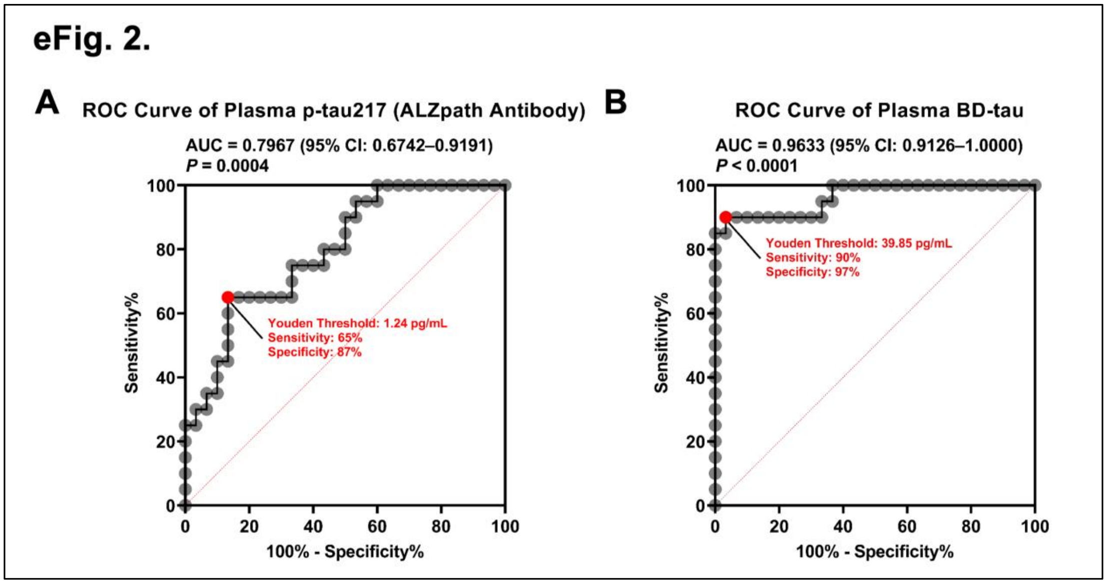


**eFig. 2. Derivation of study-specific plasma p-tau217 and BD-tau thresholds.**

**A**. The receiver operating characteristic (ROC) curve of plasma p-tau217, measured using the ALZpath antibody, was generated to assess the ability of plasma p-tau217 to discriminate Alzheimer's disease patients from population reference participants. Youden threshold value of plasma p-tau217 was identified as 1.24 pg/mL with a sensitivity of 65% and a specificity of 87%. **B**. The ROC curve of plasma BD-tau, measured using the BD-tau antibody, was generated to assess the ability of plasma BD-tau to discriminate Alzheimer's disease patients from population reference participants. Youden threshold value of plasma BD-tau was identified as 39.85 pg/mL with a sensitivity of 90% and a specificity of 97%.

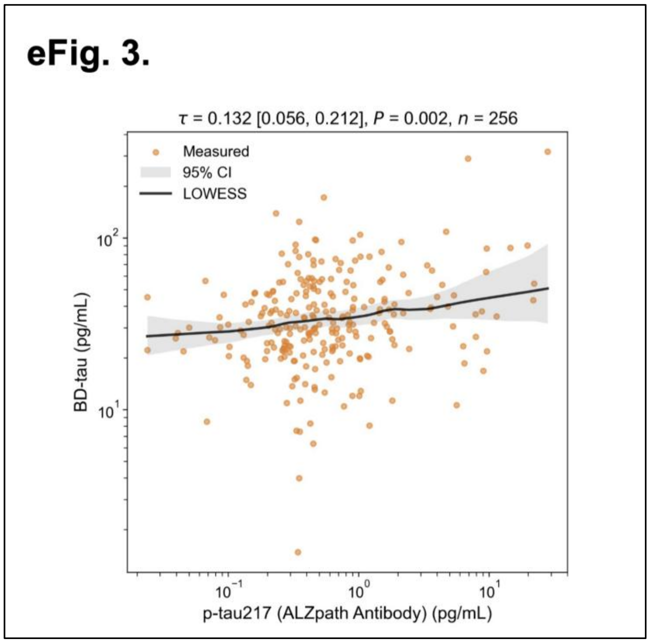


**eFig. 3. Association between plasma p-tau217 and BD-tau concentrations in blood donors.**

There is a weak association between plasma p-tau217 (measured using the ALZpath antibody) and BD-tau concentrations in blood donors. Each point represents one donor sample. Solid black curves show locally weighted scatterplot smoothing (LOWESS) fits, and gray shaded bands indicate the corresponding 95% confidence intervals. LOWESS curves are provided to visualize trends; correlation coefficients and $P$ values were calculated separately using Kendall's $\tau$. The $\tau$ value and $P$ value represent the association. p-tau217: tau protein phosphorylated at threonine 217; BD-tau: brain-derived tau.

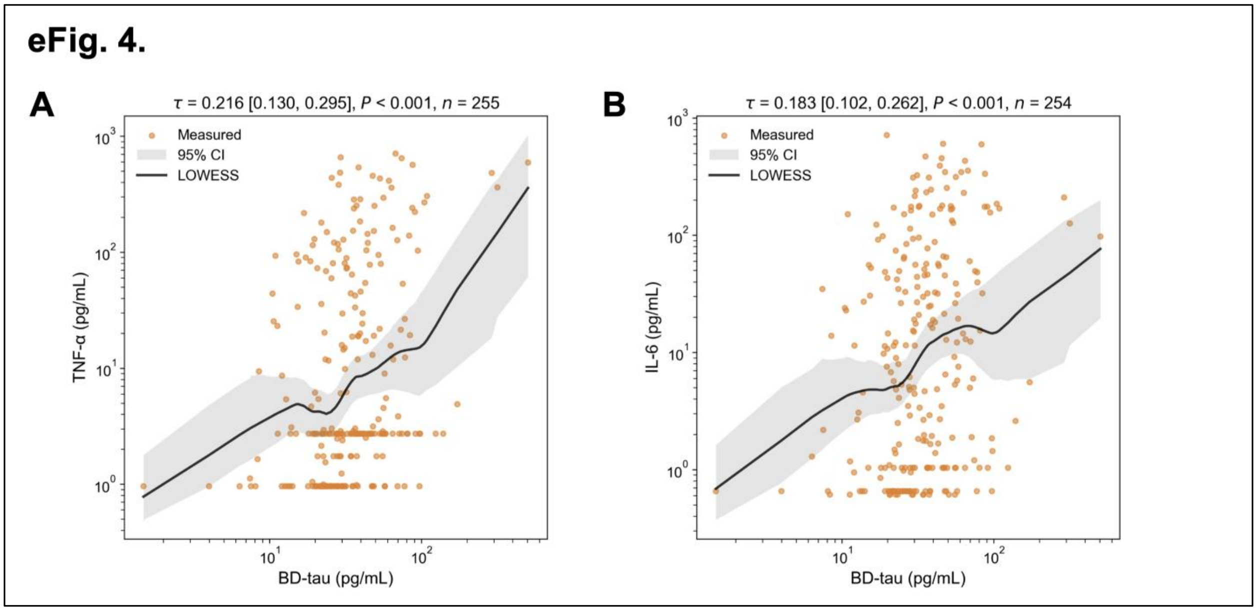


**eFig. 4. Associations between plasma BD-tau and TNF-α or IL-6 in blood donors.**

**A**. There is a weak association between plasma BD-tau and TNF-α concentrations in blood donors. **B**. There is a weak association between plasma BD-tau and IL-6 concentrations in blood donors. Each point represents one donor sample. Solid black curves show locally weighted scatterplot smoothing (LOWESS) fits, and gray shaded bands indicate the corresponding 95% confidence intervals. LOWESS curves are provided to visualize trends; correlation coefficients and $P$ values were calculated separately using Kendall's $\tau$. The $\tau$ values and $P$ values represent the associations. BD-tau: brain-derived tau; TNF-α: tumor necrosis factor α; IL-6: interleukin 6.

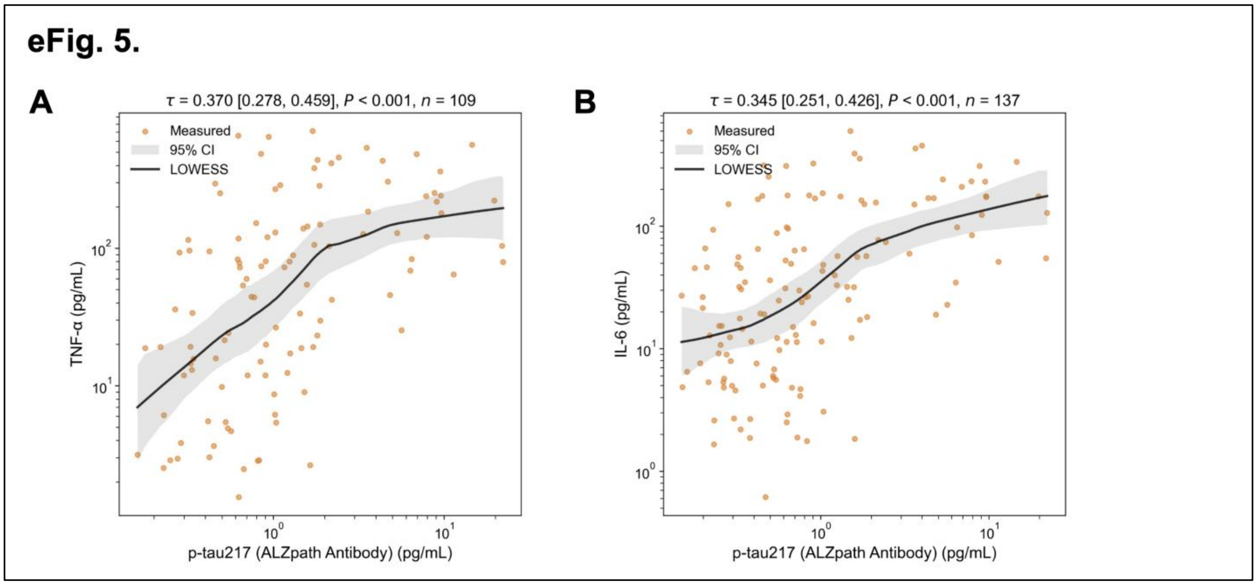


**eFig. 5. Sensitivity analyses of associations between plasma p-tau217 (ALZpath) and inflammatory biomarkers.**

**A**. Association between plasma p-tau217 (measured using the ALZpath antibody) and TNF-α concentrations in blood donors after restricting the analysis to samples without assay failures, technical outliers, or values outside the quantifiable range in the measurement of p-tau217 or TNF-α. **B**. Association between plasma p-tau217 (measured using the ALZpath antibody) and IL-6 concentrations in blood donors after restricting the analysis to samples without assay failures, technical outliers, or values outside the quantifiable range in the measurement of p-tau217 or IL-6. Each point represents one donor sample. Solid black curves show locally weighted scatterplot smoothing (LOWESS) fits, and gray shaded bands indicate the corresponding 95% confidence intervals. LOWESS curves are provided to visualize trends; correlation coefficients and *P* values were calculated separately using Kendall's *τ*. The *τ* values and *P* values represent the associations. p-tau217: tau protein phosphorylated at threonine 217; TNF-α: tumor necrosis factor α; IL-6: interleukin 6.

eFig. 6.

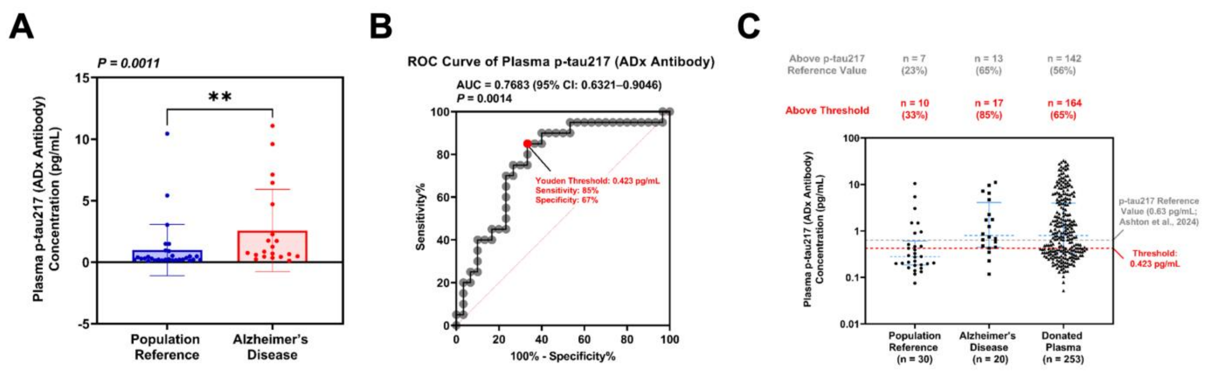


**eFig. 6. Plasma p-tau217 concentrations and threshold comparisons using the ADx antibody.**

**A**. Plasma p-tau217 concentrations were measured by using nanoneedle with a different p-tau217 antibody (ADx NeuroSciences, Ghent, Belgium) in population reference participants and Alzheimer's disease patients. Error bars indicate standard deviation. **B**. The ROC curve of plasma p-tau217, measured using the ADx antibody, was generated to assess the ability of plasma p-tau217 to discriminate Alzheimer's disease patients from population reference participants. Youden threshold value of plasma p-tau217 was identified as 0.423 pg/mL with a sensitivity of 85% and a specificity of 67%. **C**. Plasma p-tau217 concentrations in blood donors were measured using nanoneedle assays with the ADx antibody. The gray dashed line represents the prespecified published reference threshold derived from Ashton et al, and the red dashed line represents the study-specific ROC-derived Youden threshold established using population reference participants and patients with Alzheimer's disease. The numbers of blood donors with plasma p-tau217 concentrations above and below each threshold are shown. p-tau217: tau protein phosphorylated at threonine 217.

### eFig. 7.

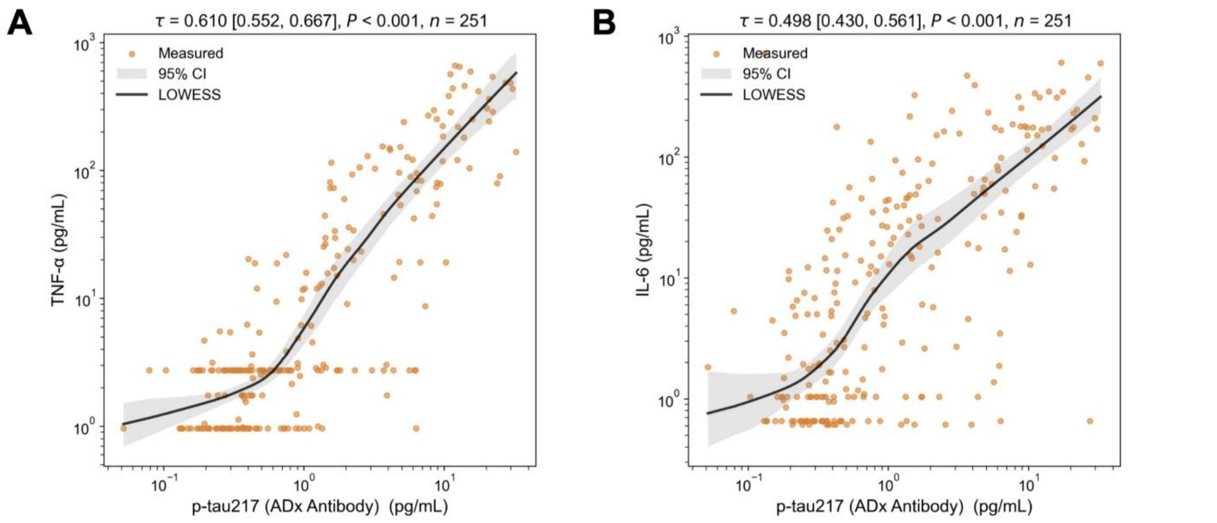


**eFig. 7. Associations between plasma p-tau217 and TNF-α or IL-6 in blood donors using a different antibody (ADx).**

**A**. Association between plasma p-tau217 (measured using the ADx antibody) and TNF-α concentrations in blood donors. **B**. Association between plasma p-tau217 (measured using the ADx antibody) and IL-6 concentrations in blood donors. Each point represents one donor sample. Solid black curves show locally weighted scatterplot smoothing (LOWESS) fits, and gray shaded bands indicate the corresponding 95% confidence intervals. LOWESS curves are provided to visualize trends; correlation coefficients and *P* values were calculated separately using Kendall's $\tau$. The $\tau$ values and *P* values represent the associations. p-tau217: tau protein phosphorylated at threonine 217; TNF-α: tumor necrosis factor α; IL-6: interleukin 6.

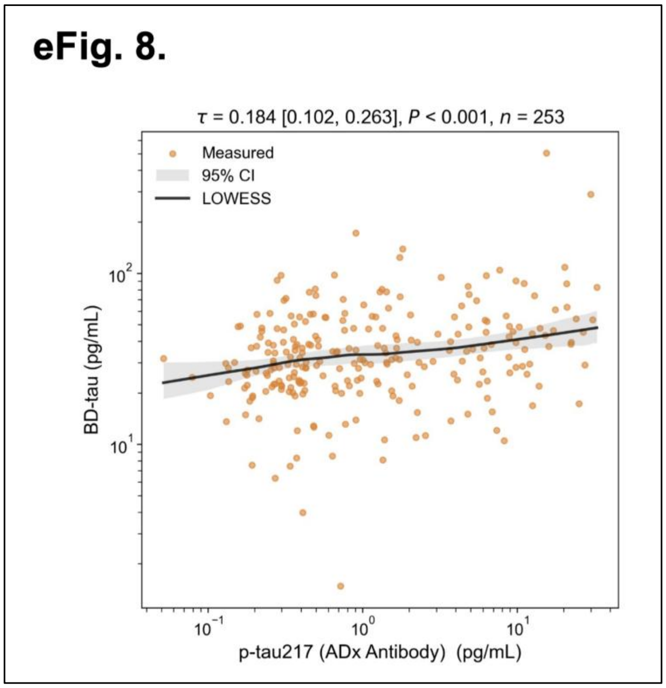


**eFig. 8. Association between plasma p-tau217 and BD-tau using a different antibody (ADx).**

There is a weak association between plasma p-tau217 (measured using the ADx antibody) and BD-tau concentrations in blood donors. Kendall's $\tau$ correlation was used to analyze the data and demonstrate the association. Each point represents one donor sample. Solid black curves show locally weighted scatterplot smoothing (LOWESS) fits, and gray shaded bands indicate the corresponding 95% confidence intervals. LOWESS curves are provided to visualize trends; correlation coefficients and *P* values were calculated separately using Kendall's $\tau$. The $\tau$ value and *P* value represent the association. p-tau217: Tau protein phosphorylated at threonine 217; BD-tau: brain-derived tau; LoD: Limit of Detection; and ULOQ: Upper Limit of Quantitation.

eFig. 9.

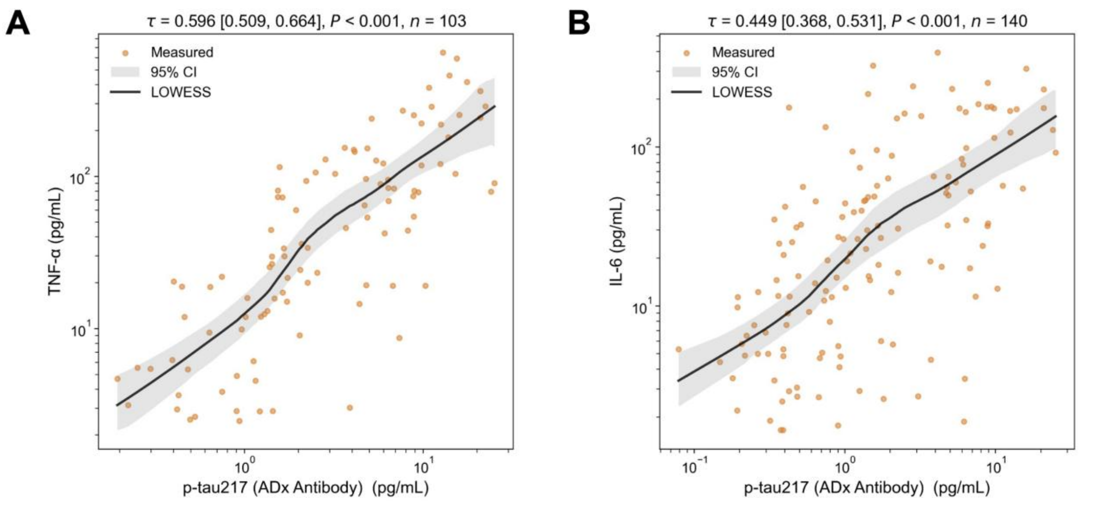


**eFig. 9. Sensitivity analyses of associations between plasma p-tau217 (ADx) and inflammatory biomarkers.**

**A**. Association between plasma p-tau217 (measured using the ADx antibody) and TNF-α concentrations in blood donors after restricting the analysis to samples without assay failures, technical outliers, or values outside the quantifiable range in the measurement of p-tau217 or TNF-α. **B**. Association between plasma p-tau217 (measured using the ADx antibody) and IL-6 concentrations in blood donors after restricting the analysis to samples without assay failures, technical outliers, or values outside the quantifiable range in the measurement of p-tau217 or IL-6. Each point represents one donor sample. Solid black curves show locally weighted scatterplot smoothing (LOWESS) fits, and gray shaded bands indicate the corresponding 95% confidence intervals. LOWESS curves are provided to visualize trends; correlation coefficients and *P* values were calculated separately using Kendall's $\tau$. The $\tau$ values and *P* values represent the associations. p-tau217: tau protein phosphorylated at threonine 217; TNF-α: tumor necrosis factor α; IL-6: interleukin 6.

**eFig. 10.**

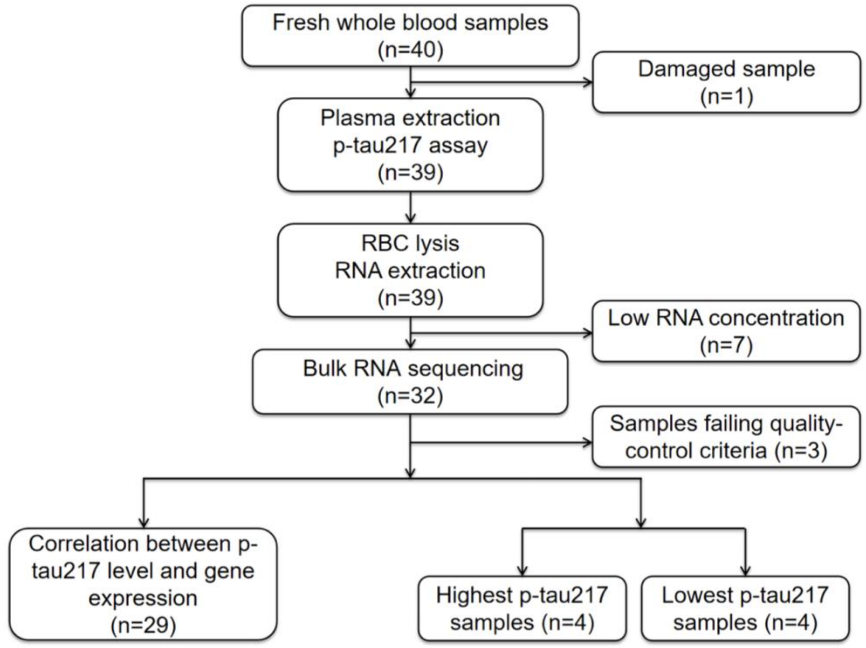


**eFig. 10. Flowchart of fresh whole blood sample processing for plasma p-tau217 measurement and bulk RNA sequencing.** Forty fresh whole blood samples were collected. One damaged sample was excluded before plasma p-tau217 analysis. Following RNA extraction, seven samples with low RNA concentration and three samples failing quality-control criteria were excluded. The remaining samples were used for correlation analysis between plasma p-tau217 levels and gene expression (n = 29). Finally, the top 4 and bottom 4 p-tau217 groups were selected for differential gene expression analysis.

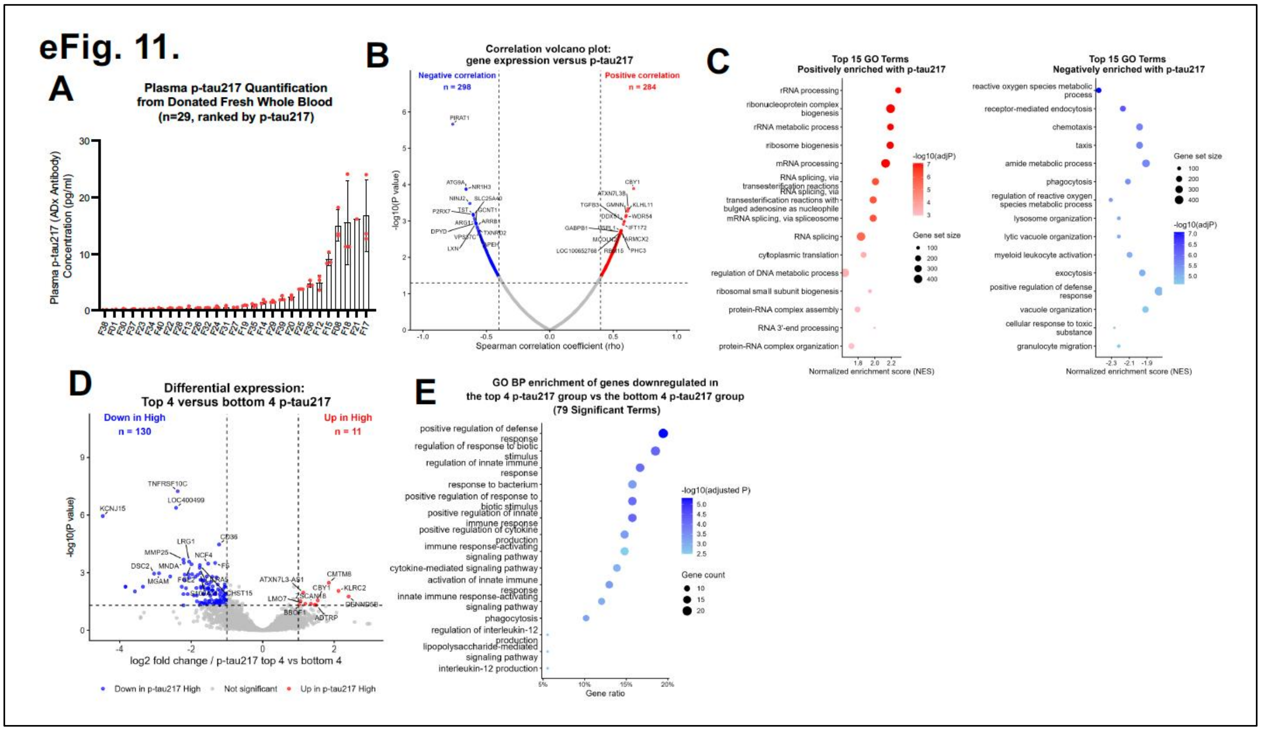


**eFig. 11. Exploratory associations between blood-cell gene expression and plasma p-tau217 measured using the ADx antibody.**

**A.** The concentrations of p-tau217 in the plasma of fresh whole blood samples obtained from 29 blood donors were detected by using ADx antibody. Error bars indicate standard deviation. **B.** Correlation volcano plot showing associations between whole blood gene expression and plasma p-tau217 levels. Spearman correlation analysis was performed between variance-stabilized gene expression values and p-tau217 levels across 29 samples. Each point represents one gene. The x-axis indicates the Spearman correlation coefficient (rho), and the y-axis shows −log10 (*P* value). In an exploratory analysis, 582 genes met the prespecified criteria of |ρ| >0.4 and nominal *P* < .05, including 284 positively and 298 negatively correlated genes. Red points indicate positively correlated genes, blue points indicate negatively correlated genes, and gray points indicate genes that did not meet the exploratory selection criteria. The top 15 genes in each direction, ranked by *P* value, are labeled. Dashed vertical lines indicate rho = ±0.4, and the dashed horizontal line indicates *P* = 0.05. **C**. Gene set enrichment analysis of GO Biological Process terms associated with p-tau217 levels measured using ADx antibody. Genes were ranked according to their Spearman correlation coefficients with p-tau217, and GSEA was performed using Gene Ontology Biological Process gene sets. The dot plots show the 15 most significant positively enriched terms (left panel) and the 11 most significant negatively enriched terms (right panel), ranked by adjusted *P* value. Dot size represents the gene set size, and dot color represents −log10(adjusted *P*), with more intense colors indicating greater statistical significance. **D.** Whole blood gene expression was compared between the top four and bottom four plasma p-tau217 levels. Differential expression analysis was performed using DESeq2. Genes with an absolute log2 fold change greater than 1 and an adjusted *P* value below 0.05 were considered differentially expressed. Red points indicate genes upregulated, blue points indicate genes downregulated and gray points indicate genes that did not meet the exploratory selection criteria in the top four p-tau217 group as compared to bottom four p-tau217 group. A total of 11 genes were upregulated and 130 genes were downregulated in the top four p-tau217 group as compared to bottom four p-tau217 group. The top 11 genes in each direction, ranked by *P* value, are labeled. Dashed vertical lines indicate log2 fold change = ±1, and the dashed horizontal line indicates an adjusted *P* value of 0.05. **E**. GO Biological Process and exploratory extreme-phenotype enrichment analysis of genes downregulated in the top four p-tau217 group compared to bottom four p-tau217 group. GO Biological Process enrichment analysis was performed on genes downregulated in the top four p-tau217 group compared with bottom four p-tau217 group. No significantly enriched GO terms were identified among the upregulated genes. A total of 79 significantly enriched terms were identified, and the 15 most significant terms ranked by adjusted *P* value are shown. Dot size represents the number of differentially expressed genes associated with each term, and dot color represents −log10 (adjusted *P* value), with darker colors indicating greater statistical significance. GO: Gene Ontology; GSEA: Gene Set Enrichment Analysis; DESeq2: Differential Expression analysis for Sequence count data.

**eFig. 12.**

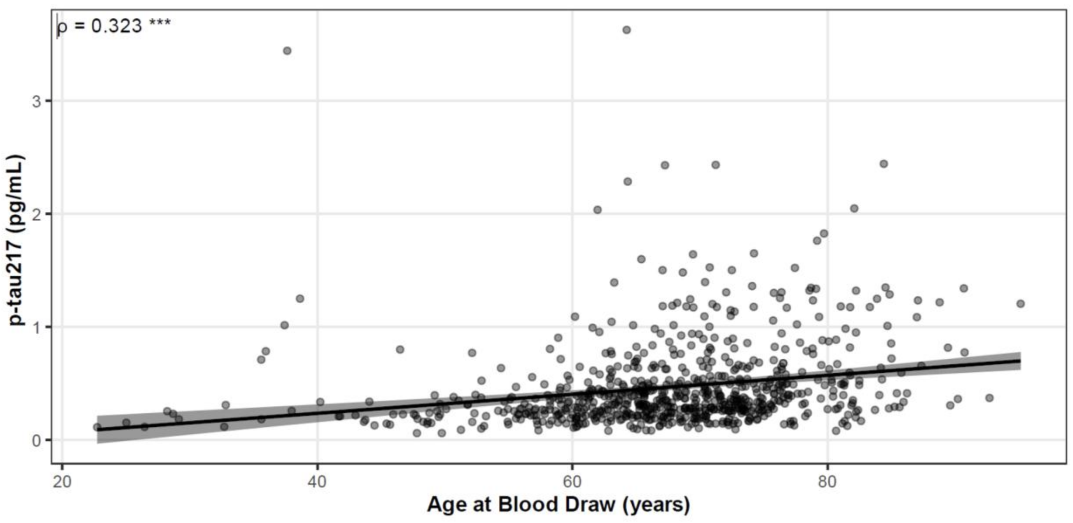


**eFig. 12. Distribution of plasma p-tau217 concentrations across age in cognitively normal individuals.**

Plasma p-tau217 concentrations were measured using the ALZpath V2 assay on the Simoa HD-X platform in PITT-ADRC participants. Each point represents one participant. Plasma p-tau217 concentrations varied across ages and were positively associated with age (Spearman ρ = 0.323). Plasma p-tau217 concentrations showed substantial interindividual variability across the adult age spectrum, with higher concentrations observed in some younger adults. This independent cohort provides descriptive context for age-related variation in p-tau217; it does not validate the nanoneedle-derived thresholds or the proportions of blood donor specimens exceeding those thresholds. Blood-donation eligibility was not assessed.

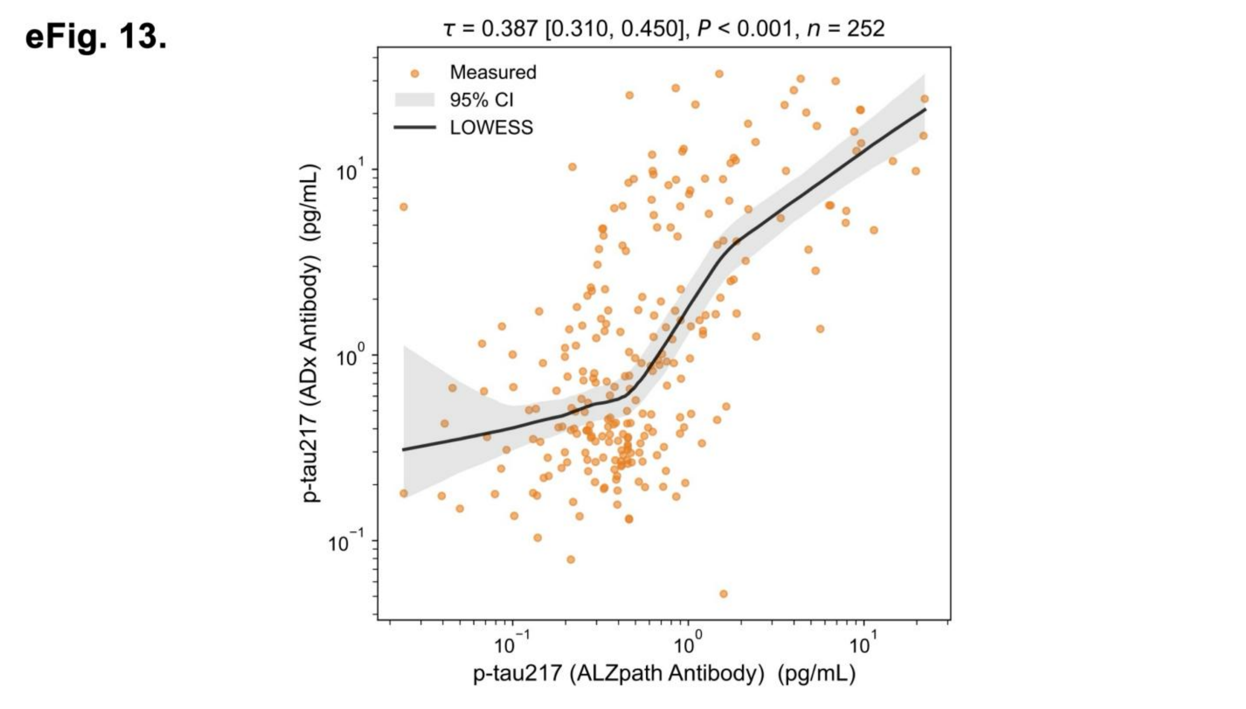


**eFig. 13. Association between plasma p-tau217 measured by ALZpath and ADx antibodies.**

There is a positive association (Kendall's $\tau = 0.387$) between the plasma p-tau217 levels, measured using the ALZpath antibody, and the plasma p-tau217 levels, measured using the ADx antibody. Each point represents one donor sample. Solid black curves show locally weighted scatterplot smoothing (LOWESS) fits, and gray shaded bands indicate the corresponding 95% confidence intervals. LOWESS curves are provided to visualize trends; correlation coefficients and *P* values were calculated separately using Kendall's $\tau$. The $\tau$ value and *P* value represent the association. p-tau217: Tau protein phosphorylated at threonine 217.

#### eTable 1. Summary of flagged replicate measurements

| Analyte/assay | Sample group | Samples | Measure number | Outliers | ULOQ | Not quantifiable | LOD |
|---|---|---|---|---|---|---|---|
| p-tau217 (ALZpath antibody) | Donor | 257 | 771 | 9 | 4 | 0 | 38 |
| | Reference | 30 | 90 | 3 | 0 | 2 | 5 |
| | AD | 20 | 60 | 0 | 0 | 0 | 0 |
| p-tau217 (ADx antibody) | Donor | 257 | 771 | 3 | 16 | 20 | 4 |
| | Reference | 30 | 90 | 2 | 0 | 0 | 1 |
| | AD | 20 | 60 | 1 | 0 | 0 | 0 |
| BD-tau | Donor | 257 | 771 | 21 | 0 | 1 | 2 |
| | Reference | 30 | 90 | 4 | 0 | 0 | 3 |
| | AD | 20 | 60 | 4 | 2 | 0 | 0 |
| TNF-α | Donor | 257 | 771 | 6 | 5 | 4 | 388 |
| IL-6 | Donor | 257 | 771 | 3 | 11 | 7 | 238 |
| p-tau217 (ALZpath antibody) | Whole blood | 29 | 87 | 5 | 0 | 0 | 3 |
| p-tau217 (ADx antibody) | Whole blood | 29 | 87 | 5 | 2 | 1 | 3 |

**Note:** Each sample was measured in triplicate, and total measurements equal three times the number of samples. Counts represent individual replicate measurements, not unique affected samples. Technical outliers, measurements above the ULOQ, and measurements reported as "not quantifiable" were excluded when calculating each sample's mean concentration. Measurements below the LoD were replaced with LoD and included in the mean. At least one retained replicate was required to calculate the mean.

**Sample groups:** Donor, plasma from blood donors; Reference, plasma from population reference participants; AD, plasma from patients with Alzheimer's disease; Whole blood, plasma prepared from fresh whole blood samples.

**Abbreviations:** AD, Alzheimer's disease; LoD, limit of detection; ULOQ, upper limit of quantification.

### eTable 2. Comparison of p-tau217 measurements and threshold exceedance between ALZpath and ADx antibodies

| Measure | ALZpath | ADx |
|---|---|---|
| Study-specific Youden threshold, pg/mL | 1.240 | 0.423 |
| Specificity at the Youden threshold, as reported | 87% | 67% |
| Donor specimens exceeding the Youden threshold, No. (%) | 53/256 (21%) | 164/253 (65%) |
| Median concentration in 252 matched samples, pg/mL | 0.455 | 0.785 |
| Samples exceeding the same exploratory cutoff of 0.63 pg/mL, No. (%) | 99/256 (39%) | 142/253 (56%) |

The referenced 0.63-pg/mL threshold [11] provides an exploratory comparison and is not a validated clinical threshold for these nanoneedle measurements.
